# Improving estimates of waning immunity rates in stochastic SIRS models with a hierarchical framework

**DOI:** 10.1101/2022.09.14.22279950

**Authors:** Punya Alahakoon, James M. McCaw, Peter G. Taylor

## Abstract

As most disease causing pathogens require transmission from an infectious individual to a susceptible individual, continued persistence of the pathogen within the population requires the replenishment of susceptibles through births, immigration, or waning immunity.

Consider the introduction of an unknown infectious disease into a fully susceptible population where it is not known how long immunity is conferred once an individual recovers from infection. If, initially, the prevalence of disease increases (that is, the infection takes off), the number of infectives will usually decrease to a low level after the first major outbreak. During this post-outbreak period, the disease dynamics may be influenced by stochastic effects and there is a non-zero probability that the epidemic will die out. Die out in this period following the first major outbreak is known as an epidemic fade-out. If the disease does not die out, the susceptible population may be replenished by the waning of immunity, and a second wave may start.

In this study, we investigate if the rate of waning immunity (and other epidemiological parameters) can be reliably estimated from multiple outbreak data, in which some outbreaks display epidemic fade-out and others do not. We generated synthetic outbreak data from independent simulations of stochastic *SIRS* models in multiple communities. Some outbreaks faded-out and some did not. We conducted Bayesian parameter estimation under two alternative approaches: independently on each outbreak and under a hierarchical framework. When conducting independent estimation, the waning immunity rate was poorly estimated and biased towards zero when an epidemic fade-out was observed. However, under a hierarchical approach, we obtained more accurate and precise posterior estimates for the rate of waning immunity and other epidemiological parameters. The greatest improvement in estimates was obtained for those communities in which epidemic fade-out was observed. Our findings demonstrate the feasibility and value of adopting a Bayesian hierarchical approach for parameter inference for stochastic epidemic models.

## 1 Introduction

Infectious diseases do not always provide life-long or long-term protective immunity after infection (Heffernan & Keeling, 2009; Mathews, McCaw, McVernon, McBryde, & McCaw, 2007). Some common examples are pertussis (Mooi, Van Der Maas, & De Melker, 2014), seasonal influenza (Camacho & Cazelles, 2013), and the A/H3N2 epidemic that occurred on the remote island of Tristan da Cunha in 1971 (Camacho et al., 2011). Furthermore, for emerging infectious diseases, sufficient biological evidence to hypothesise that either re-infection or life-long immunity is possible is often limited, as was evident during the early stages of the recent COVID-19 pandemic (Lavine, Bjornstad, & Antia, 2021; Telenti et al., 2021).

Mathematical epidemic models rely on compartmentalisation of the population into different states that are related to the infectious disease of interest (Camacho & Cazelles, 2013; Heffernan & Keeling, 2009; Kermack & McKendrick, 1927). Deterministic epidemic models that allow for replenishment of susceptibles via re-infection, births or immigration typically display damped oscillatory behaviour (Keeling & Rohani, 2011). The simplest model for such a situation is the one that allows for re-infection, the *SIRS* model. In this model, recovered individuals have immunity that wanes resulting in them becoming susceptible again.

In contrast to a deterministic model, the number of infectives in a stochastic *SIRS* model can drop to zero. This occurs due to random effects at low disease prevalence levels. Once an outbreak avoids the initial fade-out (disease extinction during the start of the epidemic), there exists a trough following the first major outbreak (Lloyd, 2004). There is a non-zero probability that disease extinction will occur during this trough (Lloyd-Smith et al., 2005). This is known as an epidemic fade-out (Alahakoon, McCaw, & Taylor, 2022; Ballard, Bean, & Ross, 2016; Bartlett, 1960; Camacho et al., 2011; Camacho & Cazelles, 2013; Lloyd-Smith et al., 2005; Meerson & Sasorov, 2009; van Herwaarden, 1997). If epidemic fade-out does not take place, the susceptible fraction will increase due to the waning of immunity, and once the effective reproduction number is greater than one, a second wave may be started. The likelihood of the occurrence of epidemic fade-out or non-fade-out depends on the model parameters of which the outbreak is modeled Anderson and May (1992).

Here, we consider a hypothetical pathogen where there is insufficient evidence to assume that recovered individuals remain immune from infection forever. We consider outbreaks of this disease observed in small closed communities (sub-populations) during short periods of time where demographic factors may be ignored. We assume outbreaks in these sub-populations take place independently, that is, the dynamics in one sub-population do not influence those in another. This assumption would be appropriate, for example, if the sub-populations were on multiple (geographically and/or temporally separated) islands or aboard multiple ships. Due to stochastic effects and variability in the characteristics of the sub-populations, we might observe epidemic fade-out in some sub-populations and not in others. In our previous work (Alahakoon et al., 2022), we introduced a novel Approximate Bayesian Computation algorithm to estimate the parameters of a stochastic epidemic model within a hierarchical framework. We tested the epidemiological applicability of this estimation framework only in the case where there is a single unknown parameter in the model. In this study, we extend our previous estimation framework to the more realistic situation in which there are multiple unknown parameters and evaluate the performance of the algorithm through a study in which the transmission rate, infectious duration and rate of waning immunity are all estimated.

In particular, conditional on an epidemic taking off, we attempt to recover the waning immunity rate when epidemic fade-outs are observed. We conduct a simulation-based experiment where multiple outbreaks of synthetic data are generated from a stochastic *SIRS* model. Some outbreaks display fade-outs while others do not. We estimate model parameters by considering each outbreak independently as well as under a Bayesian hierarchical framework. We further consider two assumptions for the prior distributions under the Bayesian framework: one in which zero appears in the support of the prior distribution for the waning immunity rate and one where it does not. We demonstrate that when the estimation is conducted independently, the waning immunity rate is often poorly estimated, particularly when an epidemic fade-out is observed. We then show that the estimates of the waning immunity rate are improved when the estimation is carried out under a hierarchical framework. Additionally, we show that, given existing knowledge from a number of previous outbreaks, the waning immunity rate for a new and in-progress outbreak may be estimated accurately under a hierarchical framework.

## 2 Background

### 2.1 The Markovian *SIRS* model in a closed sub-population

In a well-mixed sub-population of size *N*, we will denote the number of susceptibles, infectious individuals and recovered individuals by *S*(*t*), *I*(*t*), and *R*(*t*) respectively at time *t*. An *SIRS* model is parameterised by *β*, the transmission rate, *γ*, the rate of recovery, and *μ*, the waning immunity rate. The stochastic *SIRS* system can be formulated as a continuous-time Markov chain with bi-variate states (*S*(*t*), *I*(*t*)). The model structure is illustrated in Figure 1 and the transition rates are presented in Table 1. When *μ* = 0, the dynamics of this model are identical to those of the *SIR* model.

**Table 1:** Transition rates of an *SIRS* model.

| Event | Transition | Rates |
| --- | --- | --- |
| Infection | $(s, i) \rightarrow (s - 1, i + 1)$ | $\beta si / (N - 1)$ |
| Recovery | $(s, i) \rightarrow (s, i - 1)$ | $\gamma i$ |
| Loss of immunity | $(s, i) \rightarrow (s + 1, i)$ | $\mu r, (r = N - s - i)$ |

**Figure 1:**
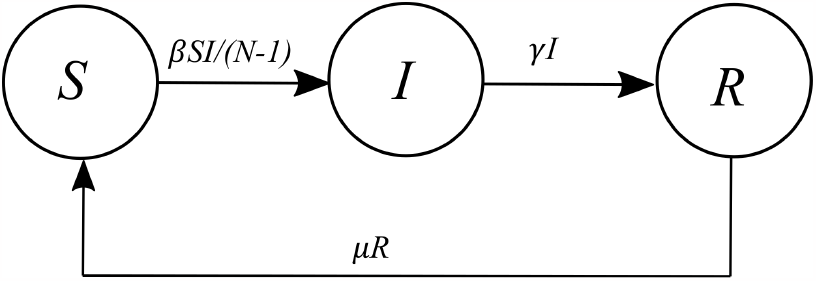
*SIRS* model structure.

In the model as we have formulated it below, in which mixing is assumed to be frequency-dependent, the constant *β* incorporates the rate at which two given individuals meet and, in the case where one is infectious and one is susceptible, transmission occurs. Alternative formulations incorporate the *N* − 1 into the constant *β* and reflect an assumption of density-dependent mixing. Since we are considering our population size to be fixed, the two formulations are equivalent in our context (Begon et al., 2002; McCallum, Barlow, & Hone, 2001).

### 2.2 A Bayesian hierarchical modelling framework

In this study, we consider the estimation of parameters under a hierarchical modeling approach. We assume that we are studying outbreaks in *K* sub-populations and each outbreak can be modeled using a continuous-time Markov chain with transition rates defined in Table 1. For *k* = 1, …, *K*, we denote the parameter set (*β*_*k*_, *γ*_*k*_, *μ*_*k*_) governing the evolution of the *k*th sub-population by ***θ***_***k***_. Under a hierarchical framework, we assume that the ***θ***_***k***_ are drawn from a common distribution (Gelman et al., 2013). Applications of hierarchical modeling frameworks to epidemiology can be found in studies such as Alahakoon, Taylor, and McCaw (2023); Cao et al. (2019); Coly, Garrido, Abrial, and Yao (2021); Lawson and Song (2010); Mathews, McBryde, McVernon, Pallaghy, and McCaw (2010).

We construct our hierarchical framework with three levels similar to that of Alahakoon et al. (2022). Level I represents the observed prevalence data ***y***_***k***_ = (*I*_*k*_(1), *I*_*k*_(2) …, *I*_*k*_(*T*_*k*_)) at *T*_*k*_ discrete time points for sub-populations *k* = 1, 2, …, *K*. Level II represents the structural relationship between the sub-population specific parameters ***θ***_*k*_ and the hyper-parameters, **Ψ**. The ***θ***_***k***_ are independent random variables with common densities *p*(***θ***|**Ψ**), which we take to be normal with mean and standard deviation given by **Ψ**. Finally, Level III represents the prior distributions for the hyper-parameters, which are generally known as hyper-prior distributions, *p*(**Ψ**) (Gelman et al., 2013).

The joint posterior density for a population consisting of *K* sub-populations is,

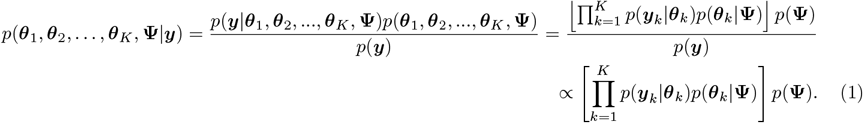

The prior distribution for the model parameters is a multivariate normal distribution with means Ψ_*β*_, Ψ_*γ*_, and Ψ_*μ*_, standard deviations *σ*_*β*_, *σ*_*γ*_, and *σ*_*μ*_, and correlations set to zero.

## 3 Materials and Methods

### 3.1 Synthetic data generation

Using the stochastic SIRS model structure that was introduced in Section 2, we constructed synthetic data for 15 sub-populations each consisting of 1000 individuals. We fixed the initial conditions of each outbreak to include one infectious person in each sub-population at the start of the outbreak. We independently generated transmission, recovery, and waning immunity rates from three truncated normal distributions. We randomly generated the transmission rates, *β*_*k*_, for the sub-populations from a normal distribution with a mean of 2.5 and standard deviation of 0.25, truncated on the interval (1, 10). We generated the recovery rates, *γ*_*k*_, from a normal distribution with a mean of 1 and a standard deviation of 0.05, truncated on the interval (0, 4). We generated the waning immunity rates, *μ*_*k*_, from a normal distribution with a mean of 0.06 and standard deviation of 0.01, truncated on the interval (0.01, 1). We used the methods of Ballard et al. (2016) to choose values for the hyper-parameters so that some of the sub-populations would display a fade-out and others would not.

See Table 2 for summary statistics of the actual values of the parameters that were generated for each of the sub-populations. Using these parameters for the *SIRS* model, we generated sample paths from the Doob-Gillespie (Doob, 1945; Gillespie, 1977) algorithm for 35 days and retained the prevalence of infections each day. If a sample path produced an initial fade-out, we discarded that sample path and repeatedly generated sample paths until an initial outbreak was observed. The criteria we used to identify an outbreak were similar to Alahakoon et al. (2022). See Supplementary Material for further details. Figure 2 shows the time-series data of the 15 sub-populations. Sub-populations 4, 8, 10, 11, 12, 14, and 15 displayed an epidemic fade-out and other sub-populations displayed multiple waves.

**Table 2:** Summary statistics of the parameters of the 15 outbreaks.

| Parameter | Mean | Standard deviation |
| --- | --- | --- |
| $\beta$ | 2.5235 | 0.3476 |
| $\gamma$ | 1.0198 | 0.0498 |
| $\mu$ | 0.0606 | 0.0073 |

**Figure 2:**
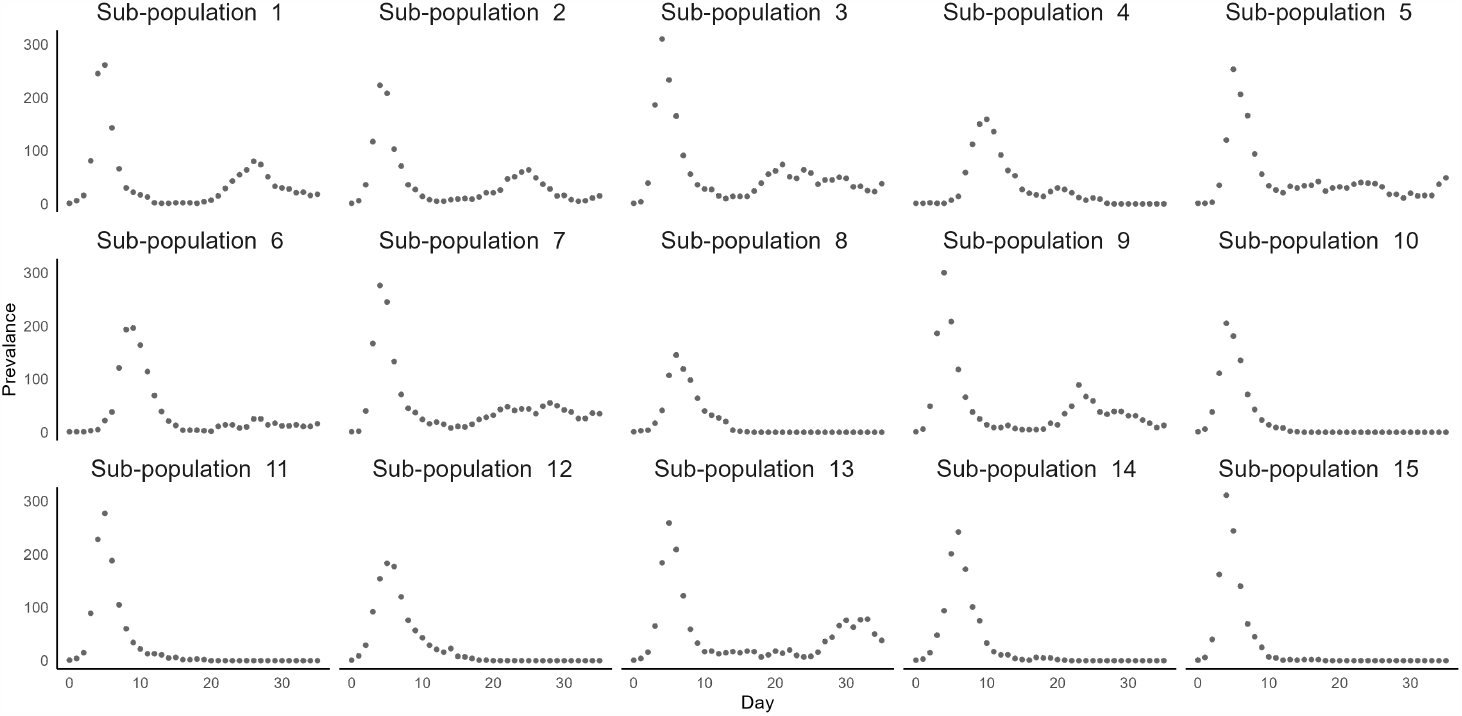
Synthetic data for fifteen sub-populations over 35 days.

### 3.2 Estimation framework

We implemented two frameworks to estimate the model parameters related to the transmission, recovery, and waning immunity rates: 1) estimation by considering each outbreak independently. 2) estimation by considering a hierarchical framework. Under each estimation framework, we made two alternative assumptions for prior distributions within the Bayesian framework.

Under the first assumption, zero is included in the support of the prior distribution for the waning immunity rate *μ* while, under the second assumption, the prior constrains *μ* to be greater than 0.01. The latter choice reflects an *a priori* assumption that immunity following infection wanes, while the first choice admits the possibility of life-long immunity following infection. Tables 3 and 4 illustrate our choice of priors for the model parameters under both assumptions.

**Table 3:** Prior distributions for parameters when the outbreaks are considered independently.

| Parameter | Assumption 1 | Assumption 2 |
| --- | --- | --- |
| $\beta$ | Uniform (0.001,10) | Uniform (0.001,10) |
| $\gamma$ | Uniform (0.00001,3) | Uniform(0.00001,3) |
| $\mu$ | Uniform (0,0.2) | Truncated Normal (0.03, 0.1<sup>2</sup>, 0.01, 0.2) |

**Table 4:** Prior distributions for hyper-parameters under the hierarchical framework.

| Hyper parameter | Assumption 1 | Assumption 2 |
| --- | --- | --- |
| $\Psi_\beta$ | Uniform (0.001, 10) | Uniform (0.001, 10) |
| $\sigma_\beta$ | Uniform (0, 2.5) | Uniform (0, 2.5) |
| $\Psi_\gamma$ | Uniform (0.00001, 3) | Uniform (0.00001, 3) |
| $\sigma_\gamma$ | Uniform (0, 1) | Uniform (0, 1) |
| $\Psi_\mu$ | Uniform (0, 0.2) | Uniform (0.01, 0.2) |
| $\sigma_\mu$ | Uniform (0, 0.15) | Uniform (0, 0.15) |

We conducted parameter estimation under both assumptions for the prior when outbreaks were considered independently with the ABC-SMC algorithm of Toni, Welch, Strelkowa, Ipsen, and Stumpf (2009). See Supplementary Material S1.1 for the details of our calibration of the algorithm. We also used the two-step algorithm of Alahakoon et al. (2022) to estimate the parameters under a stochastic hierarchical framework. See Supplementary Material S2.1 and S2.2 in relation to the calibration and diagnostics of this step. When estimating the hyper-parameters of the conditional prior distribution, the estimated correlations of the multivariate distribution were not substantial. Therefore, we used independent conditional prior distributions. See Supplementary Material S2 for further explanation.

## 4 Results

Figure 3 shows the marginal posterior distributions for the sub-population-specific waning immunity rates when parameter estimation was carried out independently for each outbreak. Irrespective of the assumed prior, the posterior distributions of the sub-populations that did not fade out had similar shapes. For the sub-populations that did fade out, the posterior distributions were strongly skewed and truncated at the lower bound (at or close to zero) of the prior distribution. This latter result for sub-populations displaying fade-out was expected: there is little if any information in the time-series for these sub-populations to inform the estimate for the rate of waning immunity, *μ*, and a value equal to or close to zero, yielding *SIR*-like dynamics, can provide a sufficient explanation for the observed data, despite the fact that the data were generated from an *SIRS* model with a waning immunity rate above zero. See Supplementary Material S1.2 for a comparison of posterior median and Highest Posterior Density (HPD) intervals (Chen, Shao, & Ibrahim, 2012) computed from *HDInterval* package in *R* (Meredith & Kruschke, 2020) and for visual diagnostics of other parameters.

**Figure 3:**
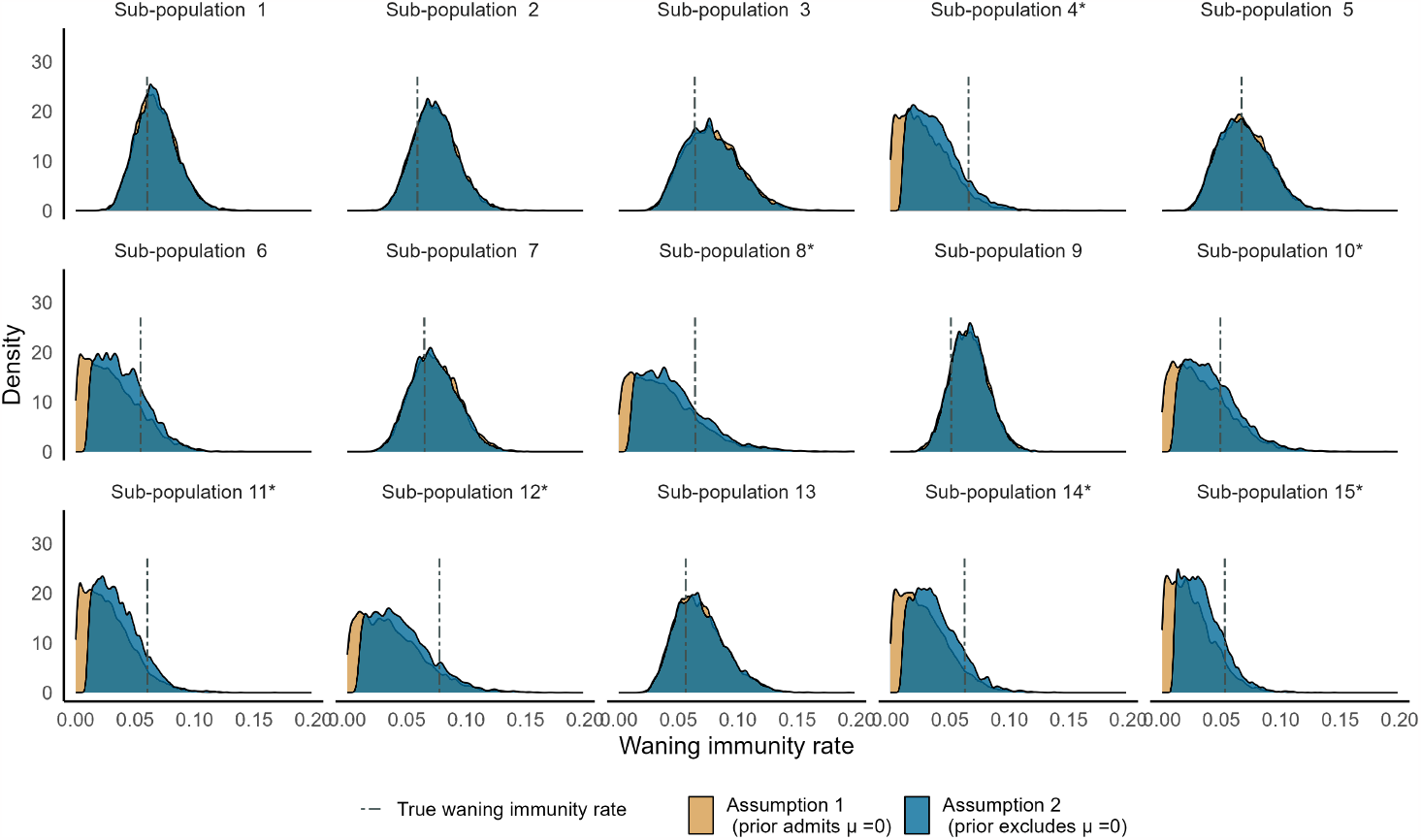
Independent estimation: Posterior distributions for the rate of waning immunity, *μ*, for the sub-populations under two assumptions for the prior. Asterisks represent sub-populations that experienced epidemic fade-out.

As we did not observe a substantial difference between the marginal posterior distributions under the different assumptions for the prior, hereafter we focus on the second assumption in which the prior for the waning immunity rate excludes zero (see Supplementary Material S2.5 for results under the alternative choice of prior). We compared estimates for model parameters under independent and hierarchical inference frameworks. Figure 4 illustrates the marginal posterior distributions for the rate of waning immunity, *μ* (plot (A)), and transmission, *β* (plot (B)). Particularly for sub-populations that displayed epidemic fade-outs, there is a striking difference between the posterior distributions for the rate of waning immunity obtained under the two frameworks. The shapes of these distributions changed from highly positively skewed and strongly biased towards zero (independent analysis) to slightly negatively skewed and with minimal bias (hierarchical analysis).

**Figure 4:**
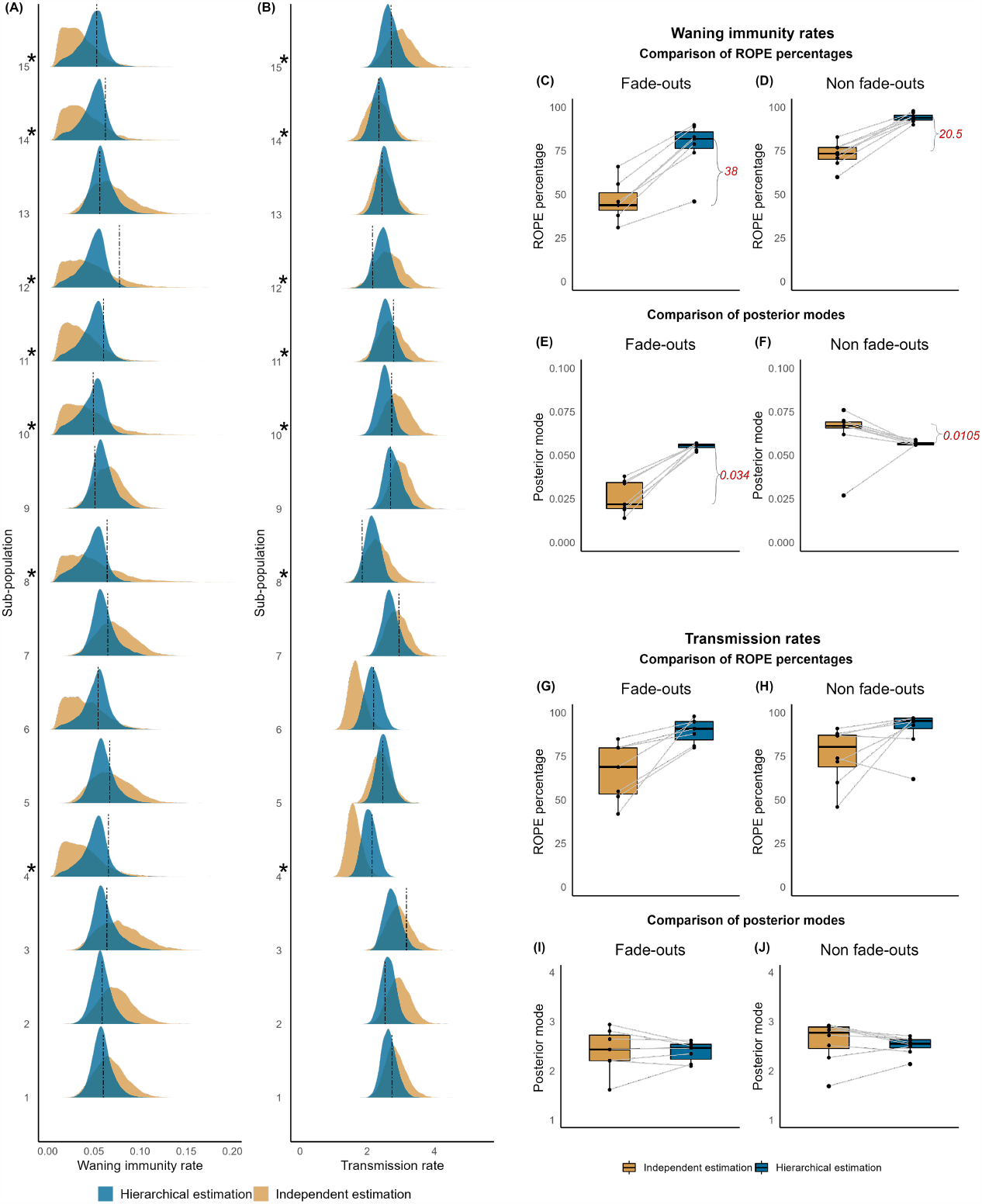
Hierarchical vs. independent estimation (assuming a prior that excludes *μ* = 0): **Left panel :** Marginal posterior distributions for waning immunity (**plot (A)**) and transmission rates (**plot (B)**). Asterisks represent sub-populations that experienced epidemic fade-out. **Top right panel :** Paired comparison of ROPE percentages (**plot (C)**) and posterior modes (**plot (D)**) with independent and hierarchical estimation frameworks for the waning immunity rate when fade-outs and non fade-outs are observed. **Bottom right panel :** Paired comparison of ROPE percentages (**plot (E)**) and posterior modes (**plot (F)**) with independent and hierarchical estimation frameworks for transmission rate when fade-outs and non fade-outs are observed.

The right panel of Figure 4 illustrates the extent of improvement of parameter estimates under a hierarchical analysis in comparison to an independent analysis. For this, we used the Region of Practical Equivalence (ROPE) criterion (J. Kruschke, 2014; J. K. Kruschke, 2013, 2018) and the posterior modes of both *μ* and *β*. We used the ROPE criterion (see Supplementary Material S2.6) to identify the percentage of the 95% Highest Posterior Density (HPD) intervals of *μ* and *β* that were included inside the ROPE. For the waning immunity rates of the sub-populations that displayed epidemic fade-outs (plot (C)), the median increase was 38%. The corresponding increase for those sub-populations that did not display fade-out (plot (D)) was 20.5%. The median of the posterior modes for *μ* increased by 0.034 (plot (E)) compared to the independent analysis for sub-populations that observed fade-outs. However, for those that did not display fade-out, the variability of posterior modes diminished (plot (F)). Overall, under a hierarchical analysis, estimates for the rate of waning immunity, *μ*, improved; and the improvement was larger for sub-populations that display epidemic fade-out.

For the ROPE percentages for *β*, the results were similar to those for *μ*. The median increase was 22% (plot (G)) and 15% (plot (H)) for sub-populations that did and did not experience fade-out respectively. The variability of the posterior modes of sub-populations that did and did not experience fade-out diminished (plots (I) and (J)) and we did not observe a shift between the medians of the posterior modes. For the recovery rates, *γ*, we made similar observations as for the transmission rates. See Supplementary Material S2.6.

Overall, in comparison to an independent analysis, a hierarchical framework dramatically improved the estimated posterior densities. Supplementary Material S2.5 provides full details for the analyses, including all marginal posterior densities under both choices for the prior on the rate of waning immunity.

Figure 5 shows the posterior distributions of the hyper-parameters under the hierarchical analysis and with a prior that excludes *μ* = 0 (assumption 2). Supplementary Material S2.7 presents estimates under the alternative assumption for the prior. Table 5 shows the posterior medians and HPD intervals for the hyper-parameters under both assumptions for the prior. The marginal posterior densities of the hyper-parameters for transmission rate and recovery rate were well estimated, with the highest posterior density for the hyper-mean lying very close to the sample-mean for the sub-population parameters. Estimates for the hyper-variances for transmission and recovery rates were also accurate and precise. For the rate of waning immunity, both the hyper-mean and hyper-standard deviation were similarly well estimated, with a noticeably improved estimate under the prior that excludes *μ* = 0 (assumption 2).

**Table 5:** Summary statistics for the posterior distributions of the hyper-parameters.

| Hyper parameter | Assumption 1<br>(prior admits $\mu = 0$) | | Assumption 2<br>(prior excludes $\mu = 0$) | |
| --- | --- | --- | --- | --- |
|  | Posterior median | HPD interval | Posterior median | HPD interval |
| $\Psi_\beta$ | 2.5144 | (2.2409, 2.7656) | 2.5573 | (2.2777, 2.7360) |
| $\sigma_\beta$ | 0.3956 | (0.1333, 0.5983) | 0.3888 | (0.1483, 0.6043) |
| $\Psi_\gamma$ | 1.0198 | (0.9280, 1.0957) | 1.0302 | (0.9412, 1.1088) |
| $\sigma_\gamma$ | 0.0539 | (0.0003, 0.1245) | 0.0506 | (0.0007, 0.1240) |
| $\Psi_\mu$ | 0.0493 | (0.0085, 0.0662) | 0.0524 | (0.0249, 0.07020) |
| $\sigma_\mu$ | 0.0143 | (0.0002, 0.0509) | 0.0095 | (0.0000, 0.0442) |

**Figure 5:**
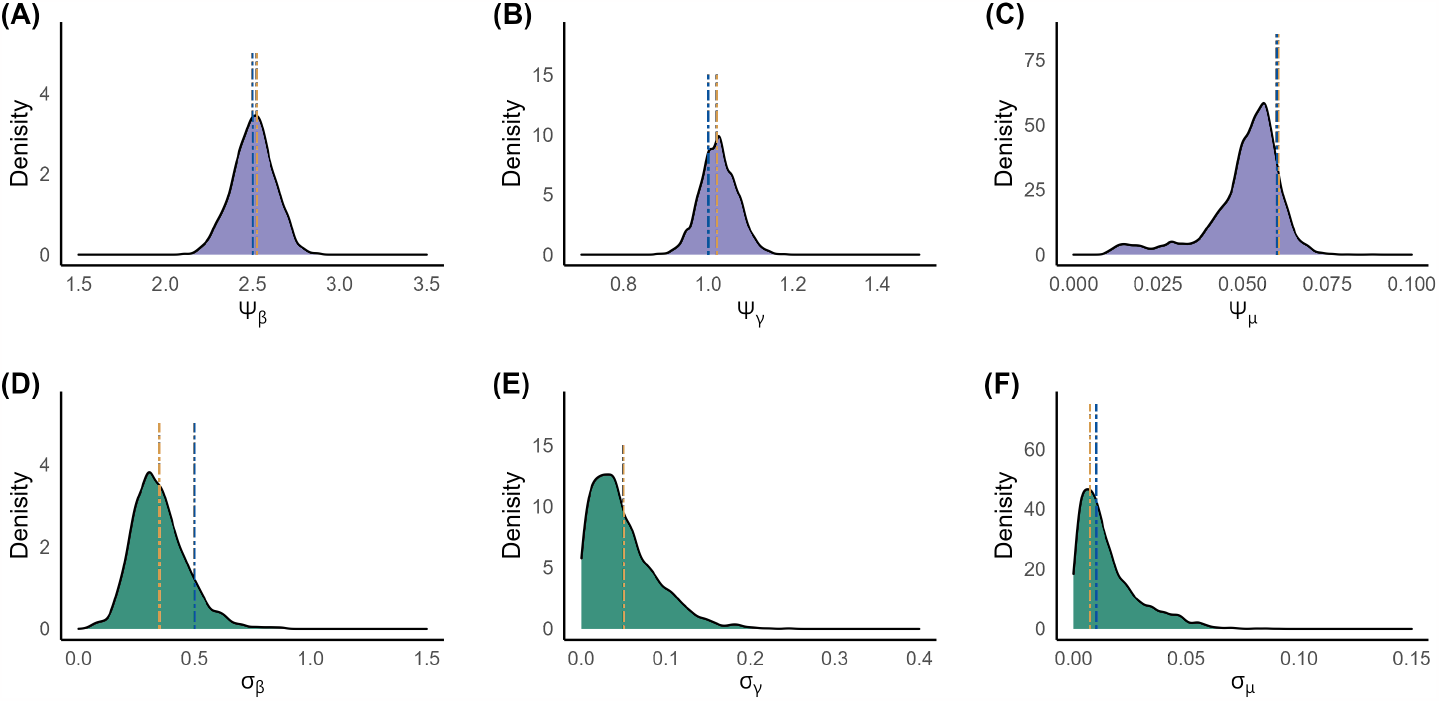
Marginal posterior distributions for the hyper-parameters for *β* (**A**), *γ* (**B**) and *μ* (**C**) under the prior that excludes *μ* = 0. **Blue dashed line** : Parameter value. **Orange dashed line :** Mean of the sub-population specific parameters.

### 4.1 Performance of the estimation framework under different parameter regimes

To evaluate the robustness of the inference framework, we generated three additional datasets of 15 sub-populations from different regions of parameter space. Given the epidemiological importance of the basic reproduction number, *R*_0_, we chose values for *R*_0_ at the hyperparametric level of 1.5, 4, and 8 to evaluate the method. Furthermore, we chose values for the hyper-parameters *ψ*_*β*_, *ψ*_*γ*_, and *ψ*_*μ*_ such that the probability of epidemic fade-out was approximately 0.5. Each of the three new datasets consisted of 15 outbreaks. Table 6 summarises the results from application of an independent and hierarchical inference approach, including those for the primary dataset in which *R*_0_ = 2.5. Full details of these additional analyses can be found in the Supplementary Material. Across all four simulation studies, the hierarchical analysis provided improved estimates for parameters at both the population (hyper-parameter) and sub-population (parameter) levels. In particular, estimates for the rate of waning immunity for those sub-populations in which fade-out was observed were notably improved, with the bias (towards an estimate of no waning) either greatly reduced or removed entirely.

**Table 6:** Performance of the hierarchical estimation method in comparison to independent estimation for four alternative epidemiological scenarios (governed by *R*_0_), using a prior that allows for *μ* = 0 (assumption 1)

| $R_0$<br>at the<br>hyperparametric<br>level | Number of<br>fade-outs<br>out of 15<br>outbreaks | Median increase in ROPE percentage<br>(from independent to hierarchical<br>estimation) | | |
| --- | --- | --- | --- | --- |
|  |  | Fade-outs | Non fade-outs | Overall increase |
| 1.5 | 9 | 21 | 38 | 29 |
| 2.5 | 7 | 36 | 20 | 34 |
| 4 | 7 | 21 | 8.5 | 27 |
| 8 | 8 | 35 | 20 | 34 |

### 4.2 Parameter estimates under incomplete time-series data

We draw the reader’s attention to the disease dynamics of sub-population 6 of our first dataset (*R*_0_ = 2.5 at the hyper-parametric level, Figure 2). This outbreak had not experienced an epidemic fade-out in 35 days (as the prevalence had not reached zero), nor had it displayed a distinct second wave. This was reflected in the posterior distribution of the waning immunity when estimated under the independent (non-hierarchical) framework, whereby the shape of the distribution was positively skewed and strongly biased towards *μ* = 0 (Figure 3). That is, *SIR*-like dynamics were not excluded. This observation motivated us to study whether a hierarchical framework, already informed by observed dynamics from other outbreaks, is able to identify the presence of waning immunity within a sub-population when only a part of the time-series (for that sub-population) is observed.

Accordingly, we drew two additional parameter sets from the same probability distributions that were used to generate the synthetic data for the 15 existing sub-populations. For each of these parameter sets we generated a new outbreak (henceforth labelled sub-populations 16 and 17). The sampled transmission rates for the two sub-populations were 2.6355 and 2.0364, the recovery rates were 1.0169 and 0.9352, and the waning immunity rates were 0.0642 and 0.0443 respectively. We first generated sample paths up to 35 days, ensuring that one of the sub-populations displayed a second wave and the other, an epidemic fade-out. We then produced an incomplete time-series data set for each sub-population by taking only the first 15 days of data. Plot (A) of Figure 6 displays the observed incomplete time-series (black solid line) and the complete time-series (black dashed line).

**Figure 6:**
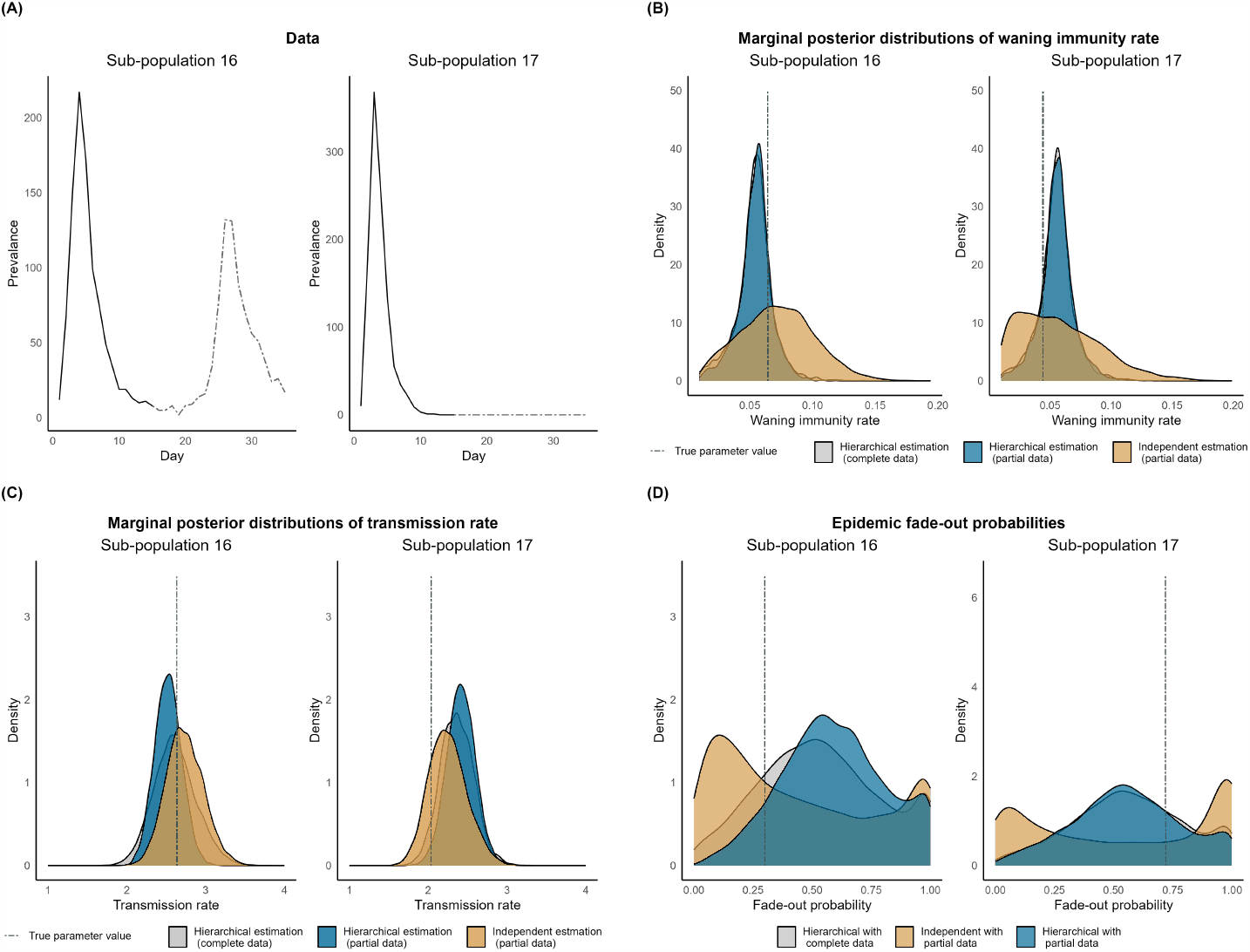
**Plot (A):** Data observed (in black line) and not yet observed (in grey dashed) for sub-populations 16 and 17. **Plot (B):** Posterior distributions for *μ* for the sub-populations 16 and 17 under parameter estimation independently and under a hierarchical framework. **Plot (C):** Posterior distributions for *β* for the sub-populations 16 and 17 under parameter estimation independently and under a hierarchical framework. **Plot (D) :** Probability of epidemic fade-out distributions from the estimated parameters. The horizontal line represents the probability fade-out calculated with the true parameter values using the methods by Ballard et al. (2016).

Under our scenario, an investigation of the first 15 sub-populations has already been conducted, from which clear evidence for waning immunity has been established, so parameter estimation for the two new sub-populations was conducted assuming a prior that excludes *μ* = 0 (that is, assumption 2 in preceding analyses). We first estimated the parameters by considering the outbreaks independently and then conducted inference under a hierarchical framework. We carried out the latter analyses by considering data from all 17 outbreaks; that is, 15 existing sub-populations with 35-day time-series data and 2 new sub-populations with 15-day time-series data.

Plots (B) and (C) of Figure 6 illustrate the posterior distributions under the two estimation frameworks for the two new outbreaks with partial data for *μ* and *β* respectively. Furthermore, for comparison, we have also plotted the posterior distributions under the hierarchical framework if complete data for all 17 populations were available. The parameter estimates for both sub-populations under the hierarchical analysis improved in comparison to those from the independent analysis, with a notable improvement in the estimate for the waning immunity rate for sub-population 17 where the strong bias towards *μ* = 0 present in the independent analysis was removed. Marginal posterior densities under the hierarchical framework with incomplete data showed only minor differences to those with complete data.

Using the methods of Ballard et al. (2016), we also calculated the epidemic fade-out probabilities, 0.3003 and 0.7199, given the true parameters for sub-populations 16 and 17 respectively. Plot (D) of Figure 6 illustrates the estimated distributions of the probability of epidemic fade-out for the two sub-populations when partial (up-to 15 days) and complete (up-to 35 days) time-series data are considered (see Supplementary Material S2.8 for details). However, neither technique (independent nor hierarchical) gave a reasonable estimate for the fade-out probability.

These additional analyses demonstrate that a hierarchical approach may support real-time analyses (where a ‘new’ outbreak is active in a new sub-population) as well as retrospective epidemiological analyses, although estimates for epidemiological parameters are likely more reliable than estimates for quantities such as the probability of epidemic fade-out.

## 5 Discussion

We have studied a hypothetical infectious disease that has a non-zero waning immunity rate. We have shown that when multiple outbreaks take place in multiple communities, parameter estimates can be expected to improve when estimation is carried out under a hierarchical framework in comparison to when the outbreaks are studied independently. Application of the parameter estimation framework introduced by Alahakoon et al. (2022) yielded improved estimates.

Epidemic fade-out is a combined result of characteristics of the sub-populations and stochastic effects at low prevalence levels. Therefore, it is possible to observe epidemic fade-out or multiple waves in different sub-populations. We have shown that when an epidemic fade-out is observed in a sub-population where multiple waves are possible, the parameter(s) that indicate the possibility of re-infection/multiple waves can be incorrectly estimated when the outbreak is studied independently. On the other hand, when a hierarchical framework is used to study multiple outbreaks, information is shared among all the sub-populations which aids in improving the estimates for re-infection (and other model parameters) even when an epidemic fade-out is observed.

Furthermore, we have shown that even when there is incomplete data for some sub-populations, the waning immunity rate can be estimated when a hierarchical framework is used.

Another possible application occurs within surveillance frameworks where data accumulation is expected. As an example, when only some of the first few outbreaks of an emerging infection display epidemic fade-out, using a hierarchical framework can aid in identifying the rate of waning immunity among those communities that observed fade-outs.

Conditional on their parameter values, we have assumed that the outbreaks evolve independently. An example of such a setting includes the outbreaks that occurred on board Australian ships during the influenza pandemic of 1918 (Alahakoon et al., 2023; Cumpston, 1919).

This work and that of Alahakoon et al. (2022) considered using hierarchical frameworks within an *SIRS* model structure. Here, we have extended the framework of Alahakoon et al. (2022) to estimate multiple parameters. We believe that this demonstration of the statistical validity of our inference method and its applicability to a foundational model in mathematical epidemiology (*SIRS* dynamics) provides a robust platform for the method to be applied to actual data. We are now applying our estimation framework to such data, including an analysis of outbreaks of pandemic influenza aboard troop ships returning to Australia in 1918 (Alahakoon et al., 2023).

The estimation framework is applicable for any process that can be described using a compartmental stochastic model, including those in infectious disease epidemiology and other areas such as within-host dynamics. For example, in clinical trial settings where pharmaco-kinetic pharmaco-dynamic models are used routinely (Cao et al., 2019; Sun, McCaw, & Cao, 2022) in combination with Bayesian hierarchical methods, there is clear evidence that stochastic effects are present and multi-wave behaviour (e.g., recrudescence of malaria within the host), as well as extinction, may be observed.

## Data Availability

All the data synthetic data and relevant details included (including the URL) can be found in the Supplementary Material.

## 6 Acknowledgements

Unless otherwise mentioned, computations were done in MATLAB or R across 32 clusters. All the computations were carried out by the use of the Nectar Research Cloud (project Infectious Diseases), a collaborative Australian research platform supported by the National Collaborative Research Infrastructure Strategy (NCRIS). All the plots were generated with ggplot2 (Wickham, 2016) in R. The codes are publicly available (see Supplementary Material for details).

