## Supplementary material for "Improving estimates of waning immunity rates in stochastic SIRS models with a hierarchical framework"

Sections S1 to S2 present calibration, diagnostics, and additional results relevant to the main dataset of the paper (that is, the dataset with $R_0 = 2.5$ at the hyper-parametric level). The analysis of the other three datasets which is introduced and presented in Section 4.1 of the paper ($R_{=}1.5, 4, 8$ at the hyperparametric level) is also similar to Sections S1 and S2. Additional details relevant to these three new datasets are included in the GitHub folder (the link is provided at the end of this document).

Section S3, discusses the criteria used to identify a major outbreak. It is relevant for all four datasets.

Section S4 presents additional diagnostic and supporting results of the datasets discussed in Section 4.1 of the paper.

#### S1 Independent parameter estimation using the ABC-SMC algortitm

##### S1.1 Calibration

We used the ABC-SMC algorithm introduced by Toni, Welch, Strelkowa, Ipsen, and Stumpf (2009) to conduct parameter estimation by considering each outbreak independently. Under both assumptions, we pre-defined the tolerance intervals across 7 generations. We used the same tolerance levels for each outbreak under two assumptions: including and excluding $\mu = 0$ for the waning immunity rate. For a sub-population, we used the observed number of infectious individuals at discrete times as the observed summary statistic. We used the Euclidean distance between the observed time-series and generated time-series data as the distance metric. To identify the suitable tolerance levels for the sub-populations, we used a basic ABC algorithm. For each sub-population, we calculated the distance metric by considering the data-only time series (i.e., when the generated sample-path results in an initial fade-out. See Alahakoon, McCaw, and Taylor (2022) for an explanation). We used these metrics in the basic ABC algorithm as well as the tolerance values for the first generation of the ABC-SMC algorithm. See Figures S1 and S5 where we have plotted parameters vs. distance metric. Accordingly, we chose tolerance for the 6 remaining generations. See Table S1 for the tolerance values we used.

*

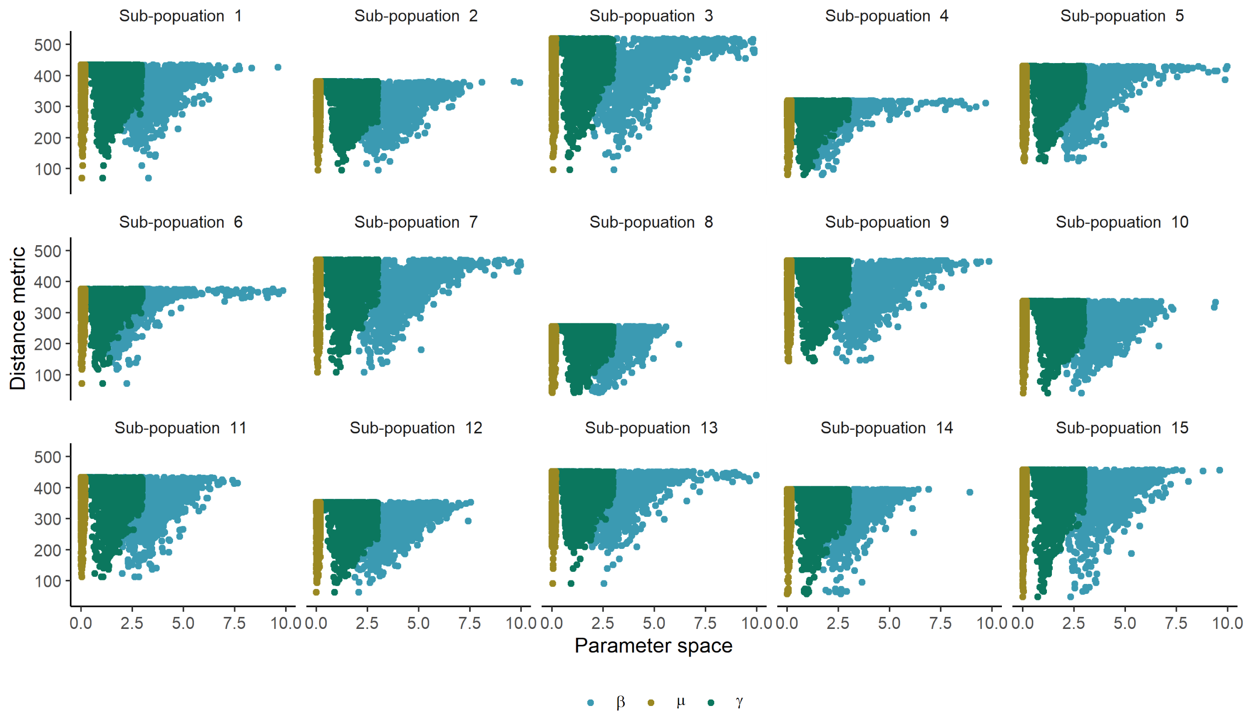


Figure S1: Scatter plots of parameter ($\beta$,$\gamma$, $\mu$) vs distance metric under assumption 1 (prior admits $\mu = 0$)for $\mu$ at $R_0 = 2.5$.

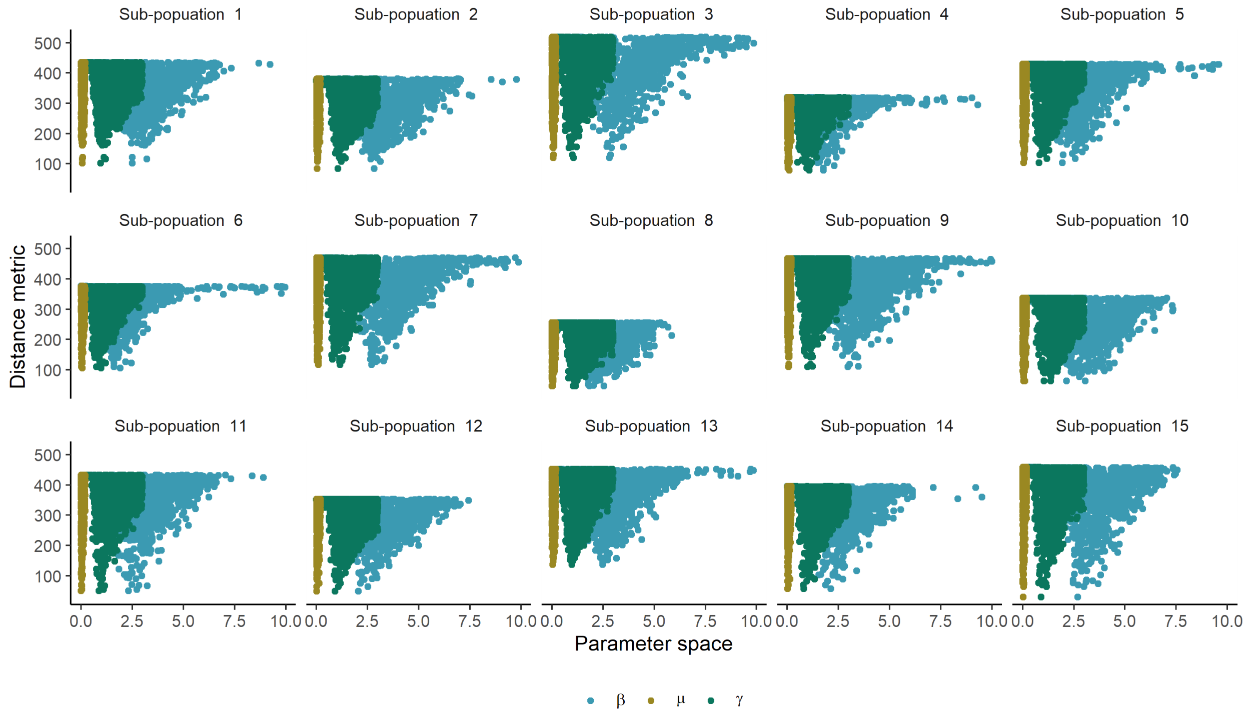


Figure S2: Scatter plots of parameter ($\beta$,$\gamma$, $\mu$) vs distance metric under assumption 2 (prior excludes $\mu = 0$) for $\mu$ at $R_0 = 2.5$.

Table S1: Pre-defined tolerance values for sub-populations under ABC-SMC for the dataset at $R_0 = 2.5$

| **Sub-population** | **Generation** | | | | | | |
|---|---|---|---|---|---|---|---|
| | **1** | **2** | **3** | **4** | **5** | **6** | **7** |
| 1 | 435 | 350 | 300 | 250 | 220 | 200 | 160 |
| 2 | 381 | 340 | 280 | 230 | 200 | 180 | 130 |
| 3 | 519 | 450 | 400 | 350 | 300 | 250 | 180 |
| 4 | 319 | 280 | 250 | 200 | 180 | 150 | 100 |
| 5 | 430 | 350 | 300 | 250 | 220 | 180 | 140 |
| 6 | 376 | 350 | 300 | 250 | 200 | 180 | 160 |
| 7 | 470 | 350 | 300 | 250 | 220 | 180 | 160 |
| 8 | 256 | 200 | 180 | 150 | 130 | 120 | 100 |
| 9 | 468 | 350 | 300 | 250 | 200 | 180 | 160 |
| 10 | 337 | 280 | 250 | 200 | 180 | 150 | 100 |
| 11 | 433 | 400 | 350 | 300 | 280 | 220 | 180 |
| 12 | 353 | 320 | 300 | 250 | 220 | 200 | 180 |
| 13 | 452 | 400 | 350 | 300 | 280 | 220 | 180 |
| 14 | 395 | 350 | 300 | 250 | 220 | 200 | 180 |
| 15 | 458 | 400 | 350 | 300 | 250 | 200 | 180 |

We obtained 5000 samples from the posterior distributions of $\beta_k$s, $\gamma_k$s, and $\mu_k$s for $k = 1, 2, \ldots, 15$ after running the ABC-SMC algorithm.

#### S1.2 Results

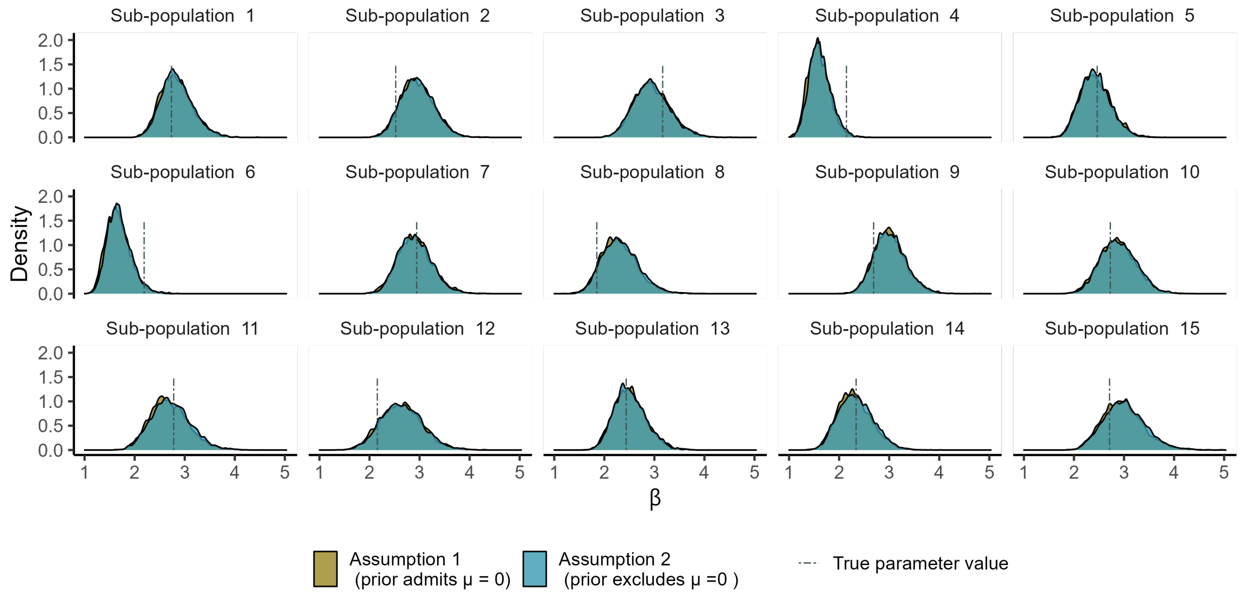


Figure S3: Posterior distributions for transmission rates using the ABC-SMC algorithm for the dataset at $R_0 = 2.5$.
These distributions visualise the posterior distributions estimated under the independent estimation framework. The distributions for each sub-population under the two assumptions; including and excluding $\mu = 0$ for prior are overlaid

Table S2: Posterior medians and HPD intervals of $\beta_k$s for the dataset at $R_0 = 2.5$

| **Sub-population** | **Parameter** | **Assumption 1 (prior admits $\mu = 0$)** | | | **Assumption 2 (prior excludes $\mu = 0$)** | | |
|---|---|---|---|---|---|---|---|
| | | **Posterior median** | **HPDI lower** | **HPDI upper** | **Posterior median** | **HPDI lower** | **HPDI upper** |
| 1 | 2.734 | 2.809 | 2.253 | 3.479 | 2.822 | 2.273 | 3.495 |
| 2 | 2.524 | 2.932 | 2.334 | 3.604 | 2.945 | 2.373 | 3.623 |
| 3 | 3.166 | 2.933 | 2.274 | 3.681 | 2.928 | 2.248 | 3.660 |
| 4 | 2.146 | 1.578 | 1.178 | 2.002 | 1.601 | 1.213 | 2.057 |
| 5 | 2.464 | 2.408 | 1.871 | 3.051 | 2.412 | 1.875 | 3.024 |
| 6 | 2.189 | 1.646 | 1.234 | 2.137 | 1.670 | 1.231 | 2.155 |
| 7 | 2.943 | 2.894 | 2.237 | 3.585 | 2.891 | 2.283 | 3.554 |
| 8 | 1.850 | 2.273 | 1.659 | 3.035 | 2.300 | 1.649 | 3.026 |
| 9 | 2.689 | 2.995 | 2.396 | 3.635 | 3.001 | 2.418 | 3.650 |
| 10 | 2.726 | 2.857 | 2.202 | 3.581 | 2.877 | 2.195 | 3.547 |
| 11 | 2.780 | 2.660 | 1.992 | 3.469 | 2.701 | 1.991 | 3.477 |
| 12 | 2.156 | 2.625 | 1.831 | 3.452 | 2.630 | 1.842 | 3.456 |
| 13 | 2.437 | 2.495 | 1.895 | 3.136 | 2.488 | 1.957 | 3.173 |
| 14 | 2.339 | 2.282 | 1.687 | 2.996 | 2.319 | 1.720 | 3.029 |
| 15 | 2.710 | 2.952 | 2.196 | 3.754 | 2.990 | 2.218 | 3.808 |

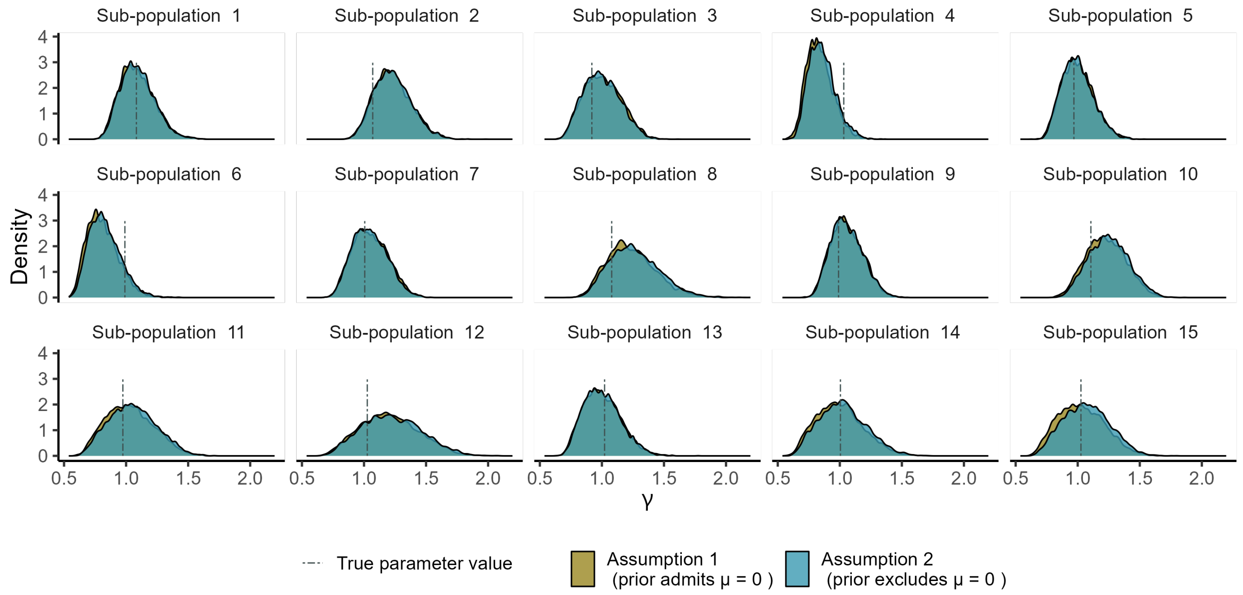


Figure S4: Posterior distributions for recovery rates using the ABC-SMC algorithm for the dataset at $R_0 = 2.5$.
These distributions visualise the posterior distributions estimated under the independent estimation framework. The distributions for each sub-population under the two assumptions; including and excluding $\mu = 0$ for prior are overlaid.

Table S3: Posterior medians and HPD intervals for $\gamma_k$s for the dataset at $R_0 = 2.5$

| | | Assumption 1 (prior admits $\mu = 0$) | | | Assumption 2 (prior excludes $\mu = 0$) | | |
|---|---|---|---|---|---|---|---|
| **Sub-population** | **Parameter** | **Posterior median** | **HPDI lower** | **HPDI upper** | **Posterior median** | **HPDI lower** | **HPDI upper** |
| 1 | 1.083 | 1.076 | 0.841 | 1.351 | 1.082 | 0.835 | 1.344 |
| 2 | 1.070 | 1.218 | 0.948 | 1.515 | 1.222 | 0.953 | 1.512 |
| 3 | 0.919 | 0.999 | 0.741 | 1.286 | 0.990 | 0.730 | 1.274 |
| 4 | 1.032 | 0.826 | 0.647 | 1.047 | 0.847 | 0.664 | 1.070 |
| 5 | 0.969 | 0.991 | 0.759 | 1.241 | 0.990 | 0.773 | 1.245 |
| 6 | 0.990 | 0.796 | 0.594 | 1.052 | 0.815 | 0.611 | 1.077 |
| 7 | 1.006 | 1.042 | 0.791 | 1.316 | 1.038 | 0.791 | 1.311 |
| 8 | 1.079 | 1.217 | 0.905 | 1.646 | 1.246 | 0.908 | 1.655 |
| 9 | 0.990 | 1.047 | 0.825 | 1.305 | 1.047 | 0.828 | 1.309 |
| 10 | 1.106 | 1.220 | 0.918 | 1.521 | 1.246 | 0.948 | 1.559 |
| 11 | 0.974 | 1.020 | 0.692 | 1.389 | 1.046 | 0.704 | 1.410 |
| 12 | 1.026 | 1.194 | 0.773 | 1.658 | 1.213 | 0.789 | 1.675 |
| 13 | 1.021 | 0.990 | 0.733 | 1.285 | 0.991 | 0.749 | 1.304 |
| 14 | 1.005 | 0.986 | 0.661 | 1.351 | 1.015 | 0.681 | 1.382 |
| 15 | 1.026 | 1.019 | 0.691 | 1.369 | 1.066 | 0.725 | 1.413 |

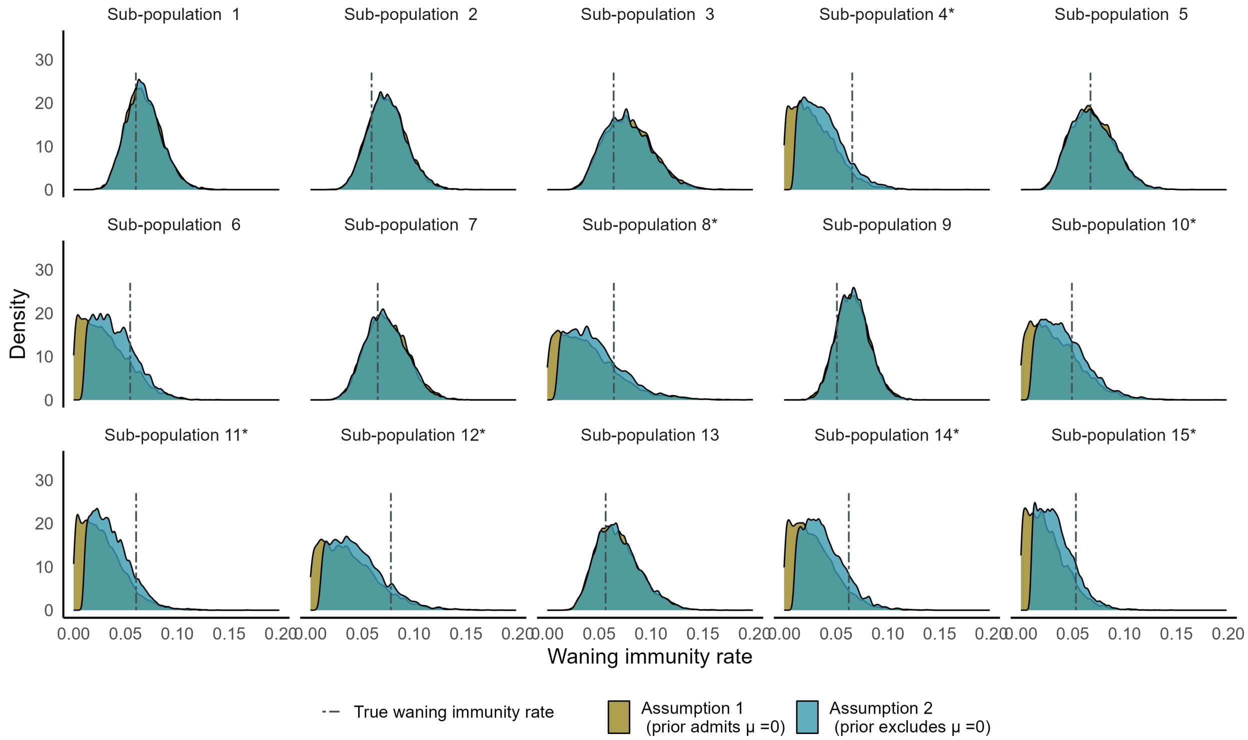


Figure S5: Posterior distributions for waning immunity rates using the ABC-SMC algorithm for the dataset at $R_0 = 2.5$. This figure is included in the manuscript (Figure 3).
These distributions visualise the posterior distributions estimated under the independent estimation framework. The distributions for each sub-population under the two assumptions; including and excluding $\mu = 0$ for prior are overlaid.

Table S4: Posterior modes, medians and HPD intervals for $\mu_k$s for the dataset at $R_0 = 2.5$. * represents the sub-populations that experienced an epidemic fade-out.

| | | **Assumption 1 (prior admits $\mu = 0$)** | | | | **Assumption 2 (prior excludes $\mu = 0$)** | | | |
|---|---|---|---|---|---|---|---|---|---|
| **Sub-population** | **Para-meter** | **Posterior mode** | **Posterior median** | **HPDI lower** | **HPDI upper** | **Posterior mode** | **Posterior median** | **HPDI lower** | **HPDI upper** |
| 1 | 0.060 | 0.021 | 0.067 | 0.036 | 0.102 | 0.020 | 0.067 | 0.036 | 0.101 |
| 2 | 0.059 | 0.018 | 0.073 | 0.040 | 0.109 | 0.021 | 0.073 | 0.039 | 0.110 |
| 3 | 0.064 | 0.020 | 0.077 | 0.038 | 0.125 | 0.024 | 0.076 | 0.035 | 0.120 |
| 4* | 0.066 | 0.000 | 0.026 | 0.000 | 0.067 | 0.010 | 0.035 | 0.010 | 0.076 |
| 5 | 0.067 | 0.005 | 0.068 | 0.031 | 0.108 | 0.015 | 0.067 | 0.030 | 0.108 |
| 6 | 0.055 | 0.000 | 0.027 | 0.000 | 0.073 | 0.010 | 0.037 | 0.010 | 0.076 |
| 7 | 0.065 | 0.020 | 0.072 | 0.036 | 0.113 | 0.017 | 0.072 | 0.037 | 0.110 |
| 8* | 0.064 | 0.000 | 0.034 | 0.000 | 0.093 | 0.010 | 0.041 | 0.010 | 0.097 |
| 9 | 0.051 | 0.023 | 0.066 | 0.036 | 0.098 | 0.018 | 0.066 | 0.038 | 0.098 |
| 10* | 0.049 | 0.000 | 0.030 | 0.000 | 0.079 | 0.010 | 0.039 | 0.010 | 0.084 |
| 11* | 0.060 | 0.000 | 0.024 | 0.000 | 0.066 | 0.010 | 0.033 | 0.010 | 0.070 |
| 12* | 0.078 | 0.000 | 0.033 | 0.000 | 0.089 | 0.010 | 0.041 | 0.010 | 0.092 |
| 13 | 0.056 | 0.018 | 0.066 | 0.030 | 0.111 | 0.020 | 0.066 | 0.032 | 0.112 |
| 14* | 0.062 | 0.000 | 0.025 | 0.000 | 0.068 | 0.010 | 0.035 | 0.010 | 0.075 |
| 15* | 0.053 | 0.000 | 0.022 | 0.000 | 0.060 | 0.010 | 0.031 | 0.010 | 0.066 |

### S2 Stochastic hierarchical parameter estimation

#### S2.1 Calibration

We used the two-step algorithm introduced by Alahakoon et al. (2022) to estimate the hyper-parameters $\Psi_\beta, \sigma_\beta, \Psi_\gamma, \sigma_\gamma, \Psi_\mu,$, and $\sigma_\mu$. According to their algorithm, the two steps are as follows:

1. Step 1: Estimating the hyper-parameters
   (a) Choose priors for the sub-population specific parameters. For each sub-population, use this prior and an ABC- based algorithm independently to get samples from the marginal posterior distributions of sub-population specific parameters.
   (b) With an approximate estimator for the likelihood at the hyper-parametric level, use the posterior samples generated in Step 1(a) to replace the likelihood function of an MCMC sampler to obtain samples from the hyper-parameters.
2. Step 2: Estimating sub-population specific parameters
   (a) Use a basic ABC algorithm to obtain samples from the posteriors of the sub-population specific parameters given the hyper-parameters (that is, sampling from the conditional prior distribution).

In this study, we used the posterior samples obtained under the two assumptions for Step 1(a). We take $\{\boldsymbol{\theta}_k^{(j)}\}_{j=1}^{N_1}$ to be size $N_1$ posterior samples of the sub-population specific parameters at the end of this step. For Step 1(b), the approximated likelihood at the hyper-parametric level is

$$\hat{p}(\boldsymbol{y}|\boldsymbol{\Psi}) \propto \prod_{k=1}^{15} \sum_{j=1}^{N_1} \frac{p(\boldsymbol{\theta}_k^{(j)}|\boldsymbol{\Psi})}{p(\boldsymbol{\theta}_k^{(j)})}, \tag{S.1}$$

where $\boldsymbol{y}$ is the observed data across the 15 sub-populations, $N$ is the size of the posterior sample of the marginal sub-population specific parameters, and $\boldsymbol{\theta}_k$ is the sub-population specific parameter $(\beta, \gamma, \mu)$. Under Assumption 2, the likelihood is of the form S.1.

Under Assumption 1, where all the sub-population specific parameters had uniform prior distributions with identical intervals for all the sub-populations, the approximate likelihood was of the form,

$$\hat{p}(\boldsymbol{y}|\boldsymbol{\psi}) \propto \prod_{k=1}^{K} \sum_{j=1}^{N_1} p(\boldsymbol{\theta}_k^{(j)}|\boldsymbol{\psi}). \tag{S.2}$$

##### S2.2 Identifying the correlation structure at the hyper-parametric level

When generating parameters for sub-populations, all the transmission, recovery, and waning immunity rates were sampled independently from normal distributions. However, in order to validate that the correlation between the hyper-parameters is not significant, under the first assumption, we estimated the hyper-parameters by assuming a correlation structure. Hence, our conditional prior distribution is a truncated multivariate normal distribution with mean $[\Psi_\beta \Psi_\gamma \Psi_\mu]$ covariance $\Sigma$ with lower bound $(0.5, 0.1, 0)$ and upper bound $(6, 2.5, 0.2)$.

We used the following prior distributions:

$$\begin{aligned}
\Psi_\beta &\sim \text{Uniform}(0.5, 6) \\
\Psi_\gamma &\sim \text{Uniform}(0.1, 2.5) \\
\Psi_\mu &\sim \text{Uniform}(0, 0.2) \\
\sigma_\beta &\sim \text{Uniform}(0, 2.5) \\
\sigma_\gamma &\sim \text{Uniform}(0, 1) \\
\sigma_\mu &\sim \text{Uniform}(0, 0.15) \\
R \sim \text{LKJcorr}(2) \quad &\text{(Prior for correlation matrix,)}
\end{aligned}$$

where,

$$R = \begin{bmatrix} 1 & \rho_{\beta\gamma} & \rho_{\beta\mu} \\ \rho_{\beta\gamma} & 1 & \rho_{\gamma\mu} \\ \rho_{\beta\mu} & \rho_{\gamma\mu} & 1 \end{bmatrix} \tag{S.3}$$

and the covariance matrix

$$\Sigma = \begin{bmatrix} \sigma_\beta & 0 & 0 \\ & \sigma_\gamma & 0 \\ 0 & 0 & \sigma_\mu \end{bmatrix} R \begin{bmatrix} \sigma_\beta & 0 & 0 \\ & \sigma_\gamma & 0 \\ 0 & 0 & \sigma_\mu \end{bmatrix} \tag{S.4}$$

We used a block-wise MCMC sampler to estimate the hyper-parameters. The programme was written in R. We used the *trialr* package in R to generate random correlation matrices from the LKJ prior. We refer to Lewandowski, Kurowicka, and Joe (2009); McElreath (2020) for more details about the use and theory related to the LKJ prior. We used the *tmvtnorm* package in R to calculate the likelihood at the hyper-parametric level when the conditional prior was a truncated multivariate normal distribution. Figure S6, shows the trace plots for $\Psi_\beta, \Psi_\gamma, \Psi_\mu, \sigma_\beta, \sigma_\gamma$, and $\sigma_\mu$. Figure S7 shows the trace plots and the posterior distributions for the correlation parameters. Figure S8 illustrates the marginal posteriors and the scatter plots between the hyper means. Although from the posterior distributions of the correlation parameters, there is no visible distinction between the posterior distribution and the corresponding prior distribution, the scatter plots between the posterior distributions of the hyper means illustrate that there is no strong correlation between the parameters.

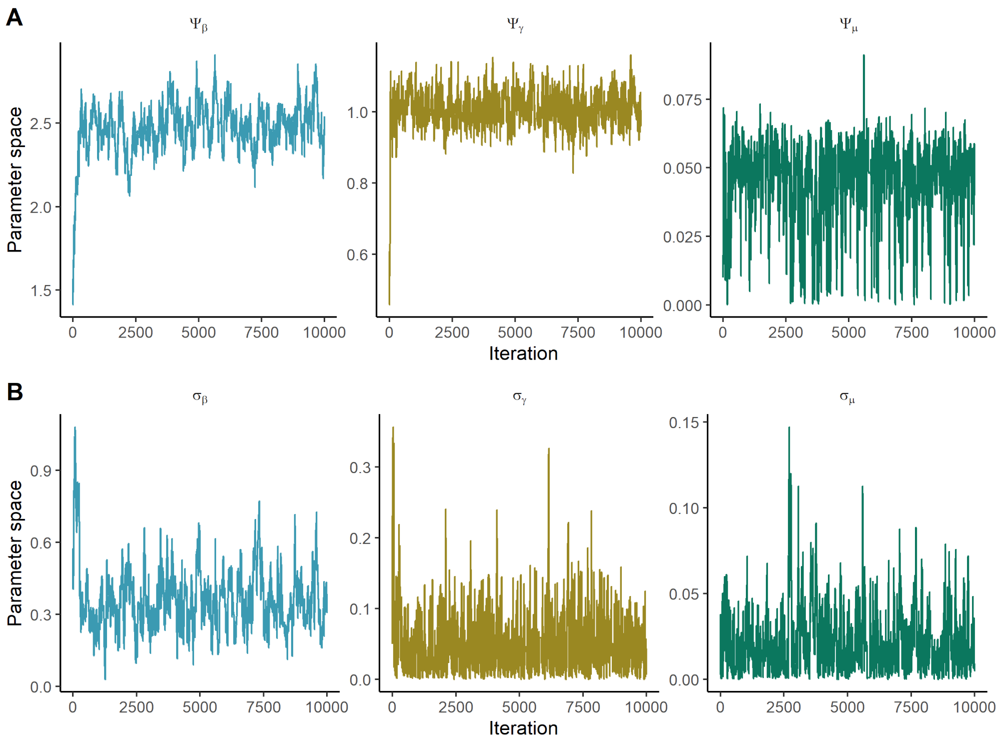


Figure S6: MCMC chains under assumption 1 (prior admits $\mu = 0$) at $R_0 = 2.5$. The MCMC algorithm was run to estimate the hyper-parameters.
**Panel A:** $\Psi_\beta, \Psi_\gamma, \Psi_\mu$. **Panel B:** $\sigma_\beta, \sigma_\gamma, \sigma_\mu$.

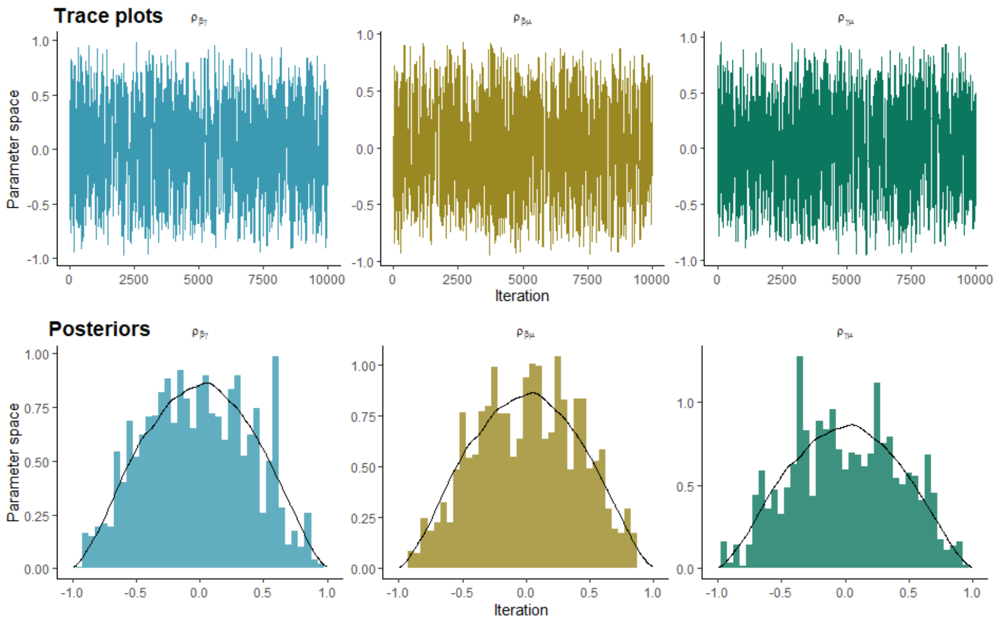


Figure S7: MCMC chains under Assumption 1 (prior admits $\mu = 0$) at $R_0 = 2.5$. The MCMC algorithm was run to estimate the hyper-parameters. **Panel A:** Trace plots for correlations. **Panel B:** Histograms of the posterior distributions for the correlation parameters. The black line represents the prior distribution.

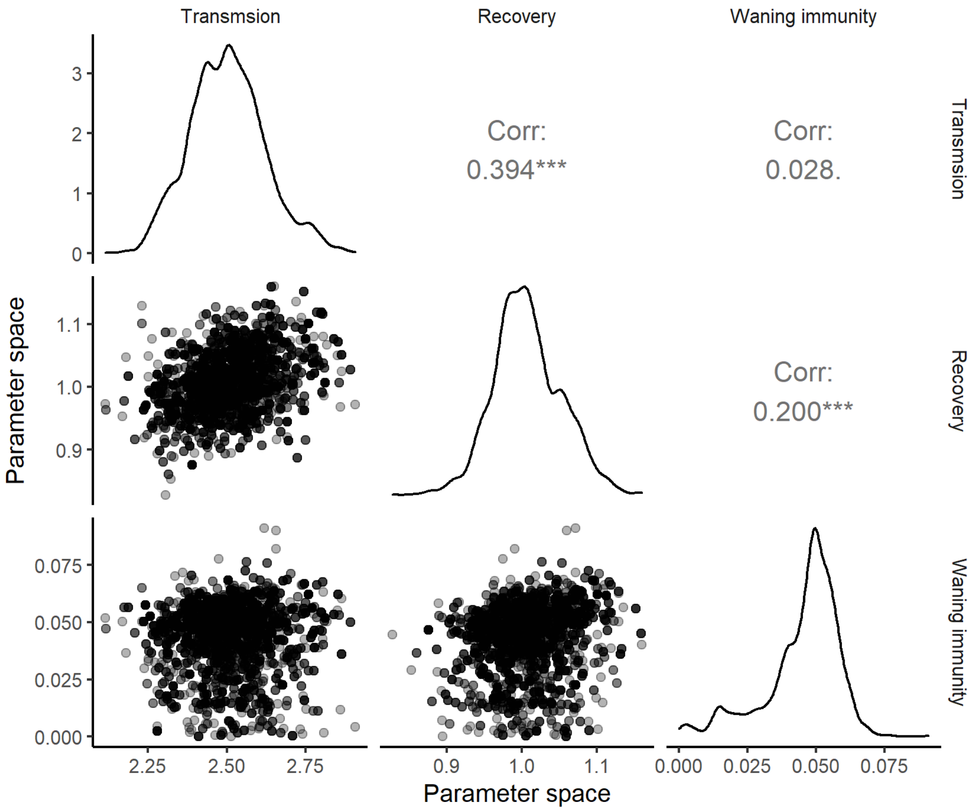


Figure S8: Marginal and bi-variate posterior distributions of $\Psi_\beta, \Psi_\gamma, \Psi_\mu$ at $R_0 = 2.5$.

##### S2.3 Estimation with independence assumption at the hyper-parametric level for the dataset at $R_0 = 2.5$

Throughout this study, as we sampled $\beta_k$s, $\gamma_k$s, and $\mu_k$s from truncated normal distributions independently, we estimated the hyper-parameters by assuming that the correlation between the hyper-parameters is zero. Accordingly, we estimated the parameters using block-wise MCMC sampling.

To proceed with block-wise sampling, we defined bi-variate normally distributed proposal distributions for $(\Psi_\beta, \sigma_\beta), (\Psi_\gamma, \sigma_\gamma)$, and $(\Psi_\mu, \sigma_\mu)$ such that for the $b$th iteration of an MCMC chain

$$\left(\Psi_{[i]}^{(b)}, \sigma_{[i]}^{(b)}\right) \sim \text{MVN}\left(\left(\Psi_{[i]}, \sigma_{[i]}\right)^{(b-1)}, \Sigma_{[i]}\right), \tag{S.5}$$

where, MVN is multivariate normal, $i = 1, 2, 3$, are $\beta, \gamma$, and $\mu$ and respectively, and $\Sigma_{[i]}$ is the variance-covariance matrix for the $i$th parameter.

Our variance-covariance matrices under both assumptions were,

$$\Sigma_{[i=1,2]} = \begin{bmatrix} 0.003 & 0 \\ 0 & 0.003 \end{bmatrix}, \quad \Sigma_{[i=3]} = \begin{bmatrix} 0.0003 & 0 \\ 0 & 0.0003 \end{bmatrix}.$$

The pseudo-code of our procedure is explained in Algorithm 1.

**Algorithm 1** Hyper-parameter $\left(\Psi_{[i]}, \sigma_{[i]}\right)$ estimation for hierarchical data

1: **Inputs:** $\{\boldsymbol{\theta}_k^{(a)}\}_{a=1}^{N_1}$, and $\boldsymbol{y}_k$ for all $k = 1, ..., K$ and $j = 1, ...., N_1$
2: **Output:** $\{\boldsymbol{\Psi}^{(b)}\}_{b=n}^{N_2}$, a sample from the marginal posteriors of the hyper-parameters after an initial burn-in period $b = 1, .., n$.
3: **Initialisation:** Define hyper-prior distributions for $p(\Psi_{[i]})$ and $p(\sigma_{[i]})$. Set initial values for the hyper-parameters as $\Psi_{[i]}^{(1)}, \sigma_{[i]}$. Set the number of iterations to run as $N_2$.
4: **for** b= 2 . . . **do**
5: **for** $i = 1:3$ **do**
6: Propose a hyper-parameters $\left(\Psi_{[i]}, \sigma_{[i]}\right)^{\text{can}}$ such that

$$\left(\Psi_{[i]}, \sigma_{[i]}\right)^{\text{can}} \sim \mathrm{Q}(\left(\Psi_{[i]}, \sigma_{[i]}\right)^{(b-1)}, \Sigma_{[i]})$$

7: Calculate the acceptance/ rejection probability $\alpha$ using $\left[\left(\Psi_{[i]}, \sigma_{[i]}\right)^{\text{can}}, \left(\Psi_{[j]}, \sigma_{[j]}\right)^{(b-1)}\right]$, $i \neq j$.
8: Generate $u \sim \text{uniform}(0, 1)$
9: **if** $\alpha > u$ **then**
10: $\left(\Psi_{[i]}, \sigma_{[i]}\right)^{\text{b}} = \left(\Psi_{[i]}, \sigma_{[i]}\right)^{\text{can}}$
11: **else**
12: $\left(\Psi_{[i]}, \sigma_{[i]}\right)^{\text{b}} = \left(\Psi_{[i]}, \sigma_{[i]}\right)^{\text{(b-1)}}$
13: Continue Step 4 until convergence can be ensured.

##### S2.4 Diagnostics: MCMC chains

From the MCMC chains, 10000 samples for each hyper-parameter were obtained. As burn-in, the first 5000 samples were discarded and the second 5000 samples were retained.

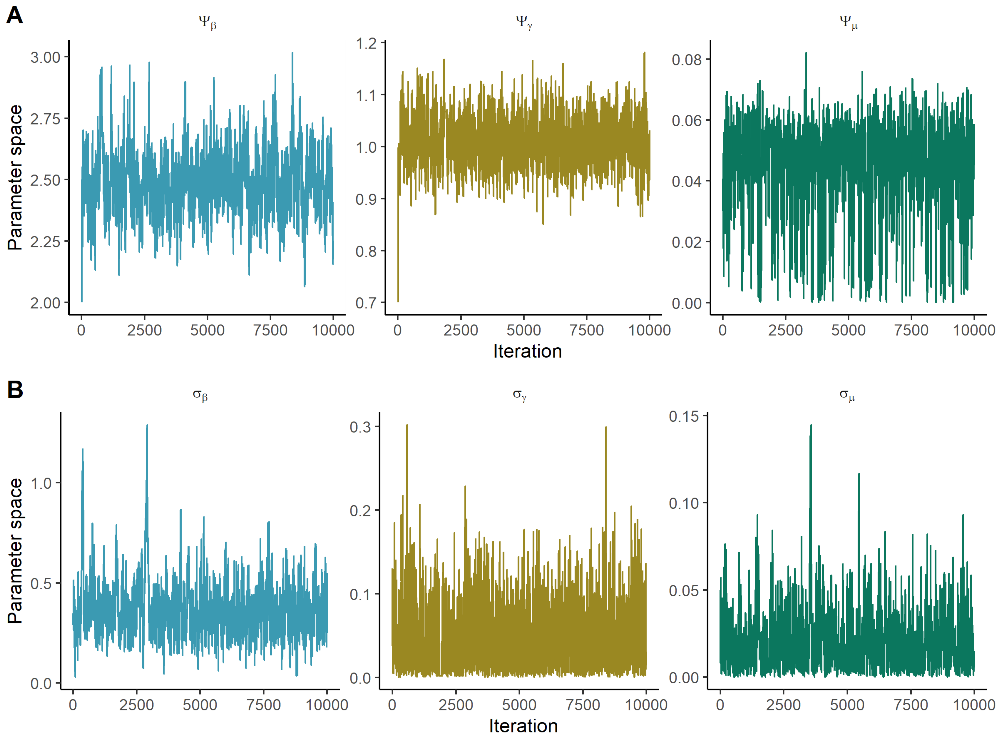


Figure S9: MCMC chains under Assumption 1 (prior admits $\mu = 0$) for the dataset at $R_0 = 2.5$ assuming that the correlation between the hyper-parameters is zero. **Panel A:** $\Psi_\beta, \Psi_\gamma, \Psi_\mu$. **Panel B:** $\sigma_\beta, \sigma_\gamma, \sigma_\mu$.

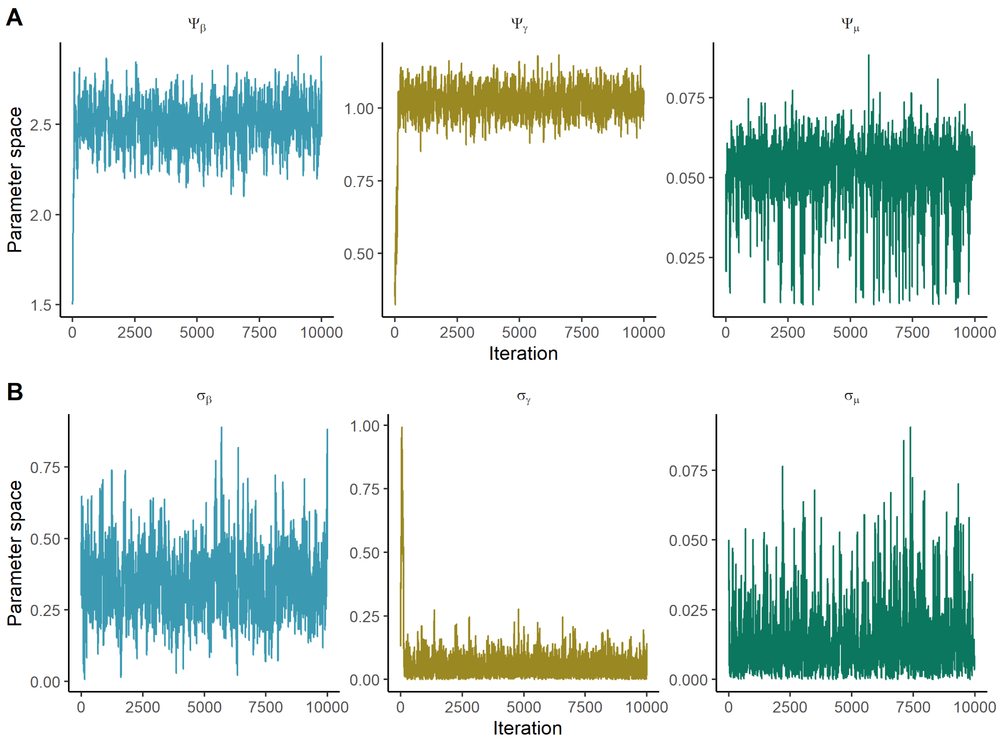


Figure S10: MCMC chains under Assumption 2 (prior excludes $\mu = 0$) for the dataset at $R_0 = 2.5$ assuming that the correlation between the hyper-parameters is zero. **Panel A:** $\Psi_\beta, \Psi_\gamma, \Psi_\mu$. **Panel B:** $\sigma_\beta, \sigma_\gamma, \sigma_\mu$.

##### S2.5 Estimation of sub-population specific parameters

Under Step 2 of the two-step algorithm, we used a basic ABC algorithm and sampled from the conditional prior, $p(\boldsymbol{\theta}_k|\Psi)$ as in the algorithm of Alahakoon et al. (2022). As tolerance levels, we used the tolerance levels of the 7th generation in the ABC-SMC algorithm for the sub-populations.

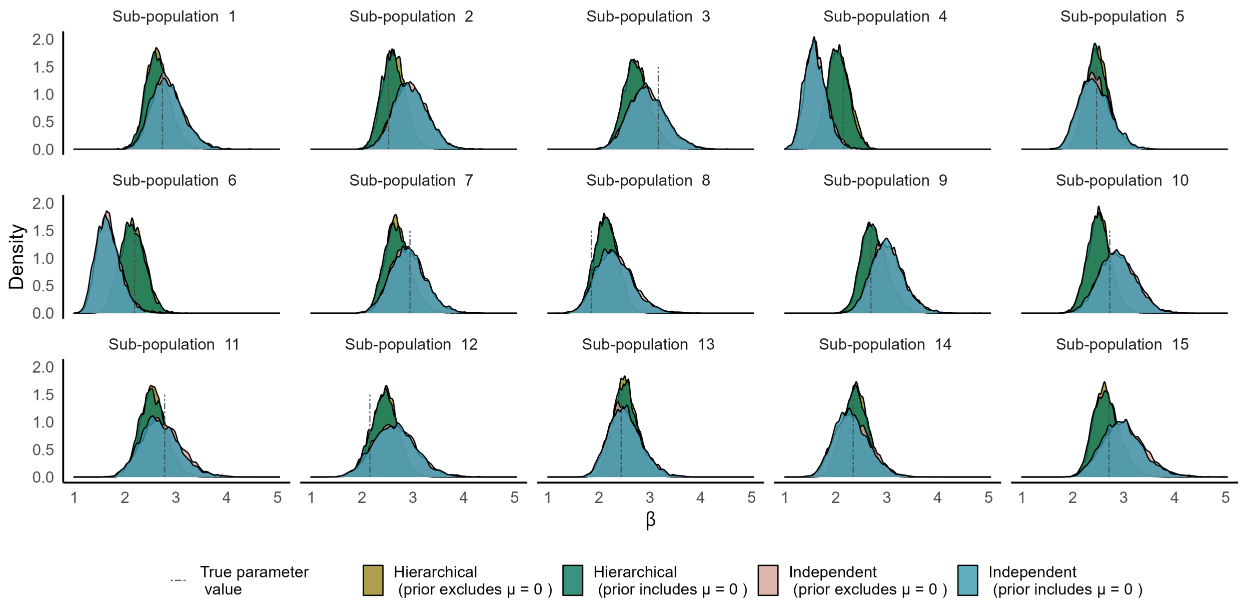


Figure S11: Posterior distributions of transmission rates under independent and hierarchical estimation frameworks for the dataset at $R_0 = 2.5$. Posterior distributions under the two assumptions with both independent and hierarchical estimation frameworks are overlaid. The blue dashed line represents the true parameter value.

Table S5: Posterior medians and HPD internals for $\beta_k$s under a hierarchical estimation for the dataset at $R_0 = 2.5$

| | | Assumption 1(prior admits $\mu = 0$) | | | assumption 2 (prior excludes $\mu = 0$) | | |
|---|---|---|---|---|---|---|---|
| **Sub-population** | **Parameter** | **Posterior median** | **HPDI lower** | **HPDI upper** | **Posterior median** | **HPDI lower** | **HPDI upper** |
| 1 | 2.734 | 2.627 | 2.170 | 3.119 | 2.638 | 2.227 | 3.118 |
| 2 | 2.524 | 2.601 | 2.181 | 3.065 | 2.626 | 2.187 | 3.053 |
| 3 | 3.166 | 2.723 | 2.270 | 3.290 | 2.738 | 2.284 | 3.240 |
| 4 | 2.146 | 2.034 | 1.615 | 2.459 | 2.058 | 1.671 | 2.493 |
| 5 | 2.464 | 2.462 | 2.033 | 2.921 | 2.489 | 2.052 | 2.931 |
| 6 | 2.189 | 2.135 | 1.680 | 2.585 | 2.158 | 1.701 | 2.607 |
| 7 | 2.943 | 2.680 | 2.200 | 3.182 | 2.682 | 2.226 | 3.223 |
| 8 | 1.850 | 2.131 | 1.708 | 2.573 | 2.149 | 1.700 | 2.571 |
| 9 | 2.689 | 2.728 | 2.287 | 3.279 | 2.745 | 2.317 | 3.284 |
| 10 | 2.726 | 2.505 | 2.073 | 2.962 | 2.515 | 2.069 | 2.938 |
| 11 | 2.780 | 2.533 | 2.056 | 3.071 | 2.555 | 2.107 | 3.063 |
| 12 | 2.156 | 2.442 | 1.910 | 2.958 | 2.451 | 1.969 | 2.985 |
| 13 | 2.437 | 2.513 | 2.074 | 2.998 | 2.524 | 2.102 | 3.039 |
| 14 | 2.339 | 2.382 | 1.873 | 2.840 | 2.392 | 1.916 | 2.873 |
| 15 | 2.710 | 2.644 | 2.157 | 3.184 | 2.647 | 2.198 | 3.195 |

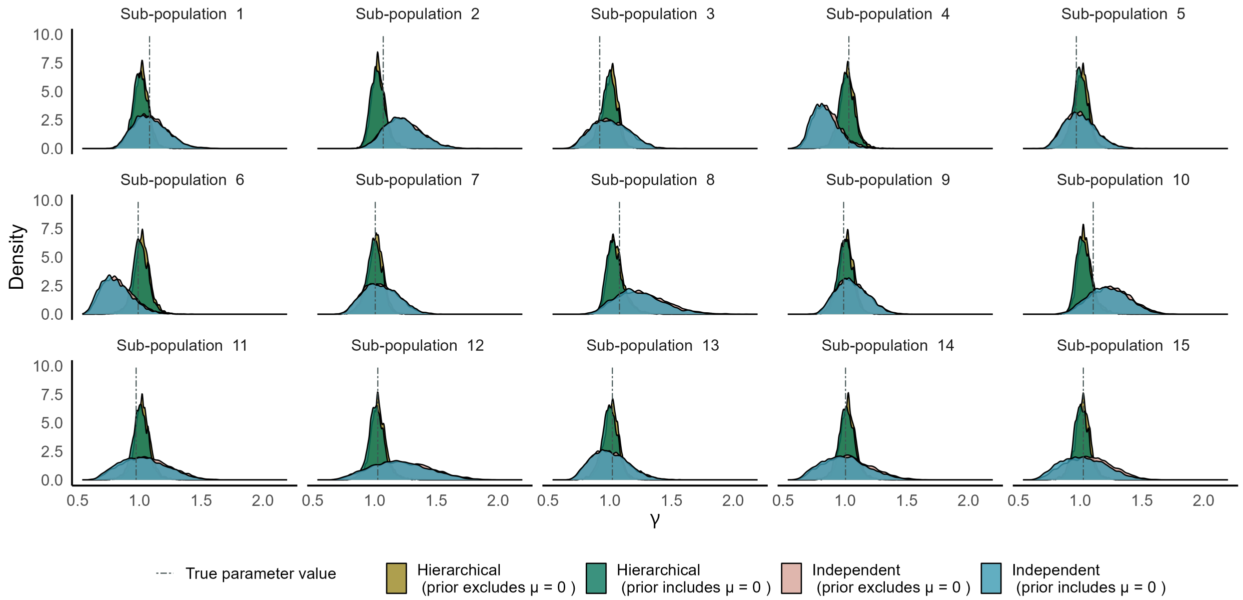


Figure S12: Posterior distributions of recovery rates under independent and hierarchical estimation for the dataset at $R_0 = 2.5$. Posterior distributions under the two assumptions with both independent and hierarchical estimation frameworks are overlaid. The blue dashed line represents the true parameter value.

Table S6: Posterior medians and HDP intervals for $\gamma_k$s under a hierarchical estimation for the dataset at $R_0 = 2.5$

| | | Assumption 1(prior admits $\mu = 0$) | | | Assumption 2 (prior excludes $\mu = 0$) | | |
|---|---|---|---|---|---|---|---|
| **Sub-population** | **Parameter** | **Posterior median** | **HPDI lower** | **HPDI upper** | **Posterior median** | **HPDI lower** | **HPDI upper** |
| 1 | 1.083 | 1.006 | 0.876 | 1.127 | 1.019 | 0.894 | 1.137 |
| 2 | 1.070 | 1.023 | 0.911 | 1.150 | 1.032 | 0.922 | 1.155 |
| 3 | 0.919 | 0.993 | 0.840 | 1.111 | 1.011 | 0.864 | 1.132 |
| 4 | 1.032 | 1.012 | 0.884 | 1.144 | 1.025 | 0.898 | 1.165 |
| 5 | 0.969 | 0.999 | 0.874 | 1.119 | 1.017 | 0.880 | 1.131 |
| 6 | 0.990 | 1.007 | 0.876 | 1.154 | 1.024 | 0.888 | 1.163 |
| 7 | 1.006 | 0.999 | 0.862 | 1.126 | 1.014 | 0.875 | 1.138 |
| 8 | 1.079 | 1.033 | 0.917 | 1.196 | 1.046 | 0.930 | 1.207 |
| 9 | 0.990 | 1.001 | 0.857 | 1.119 | 1.014 | 0.888 | 1.137 |
| 10 | 1.106 | 1.024 | 0.909 | 1.163 | 1.037 | 0.921 | 1.182 |
| 11 | 0.974 | 1.010 | 0.873 | 1.155 | 1.024 | 0.894 | 1.170 |
| 12 | 1.026 | 1.014 | 0.884 | 1.173 | 1.029 | 0.890 | 1.172 |
| 13 | 1.021 | 0.999 | 0.863 | 1.130 | 1.016 | 0.872 | 1.141 |
| 14 | 1.005 | 1.013 | 0.869 | 1.158 | 1.026 | 0.896 | 1.169 |
| 15 | 1.026 | 1.008 | 0.875 | 1.152 | 1.025 | 0.901 | 1.161 |

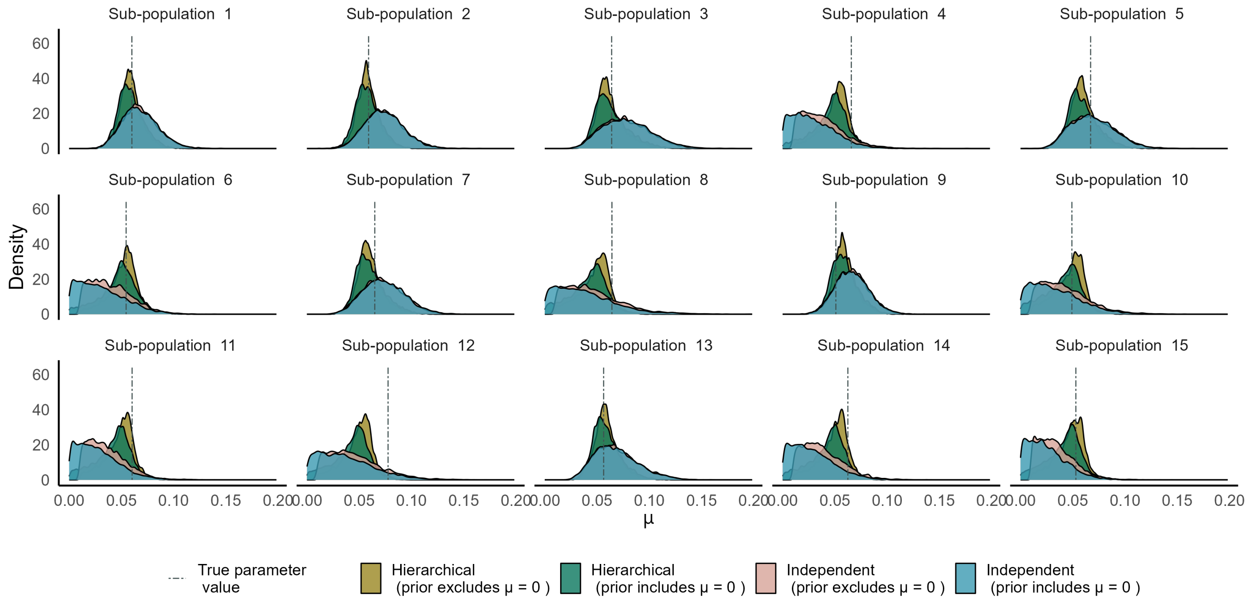


Figure S13: Posterior distributions of waning immunity rates under independent and hierarchical estimation for the dataset at $R_0 = 2.5$. Posterior distributions under the two assumptions with both independent and hierarchical estimation frameworks are overlaid. The blue dashed line represents the true parameter value.

Table S7: Posterior medians and HPD intervals for $\mu_k$s under a hierarchical estimation for the datset at $R_0 = 2.5$

| Sub-population | Parameter | Assumption 1(prior admits $\mu = 0$) | | | Assumption 2 (prior excludes $\mu = 0$) | | |
|---|---|---|---|---|---|---|---|
| | | Posterior median | HPDI lower | HPDI upper | Posterior median | HPDI lower | HPDI upper |
| 1 | 0.060 | 0.058 | 0.037 | 0.084 | 0.058 | 0.038 | 0.082 |
| 2 | 0.059 | 0.057 | 0.036 | 0.081 | 0.057 | 0.039 | 0.081 |
| 3 | 0.064 | 0.061 | 0.035 | 0.095 | 0.060 | 0.039 | 0.093 |
| 4* | 0.066 | 0.048 | 0.007 | 0.077 | 0.053 | 0.017 | 0.075 |
| 5 | 0.067 | 0.058 | 0.034 | 0.090 | 0.059 | 0.038 | 0.088 |
| 6 | 0.055 | 0.049 | 0.005 | 0.078 | 0.054 | 0.018 | 0.078 |
| 7 | 0.065 | 0.058 | 0.037 | 0.092 | 0.059 | 0.039 | 0.089 |
| 8* | 0.064 | 0.044 | 0.000 | 0.066 | 0.050 | 0.014 | 0.072 |
| 9 | 0.051 | 0.058 | 0.035 | 0.085 | 0.058 | 0.038 | 0.083 |
| 10* | 0.049 | 0.043 | 0.000 | 0.064 | 0.050 | 0.015 | 0.070 |
| 11* | 0.060 | 0.045 | 0.005 | 0.068 | 0.051 | 0.017 | 0.072 |
| 12* | 0.078 | 0.046 | 0.003 | 0.070 | 0.051 | 0.017 | 0.074 |
| 13 | 0.056 | 0.057 | 0.036 | 0.087 | 0.058 | 0.036 | 0.084 |
| 14* | 0.062 | 0.046 | 0.006 | 0.073 | 0.051 | 0.016 | 0.071 |
| 15* | 0.053 | 0.044 | 0.004 | 0.067 | 0.050 | 0.016 | 0.070 |

##### S2.6 Use of the Region of Practical Equivalence (ROPE) criterion

We defined a ROPE interval around the true parameter values such that $(\beta \pm c_1)$, $(\gamma \pm c_2)$, and $(\mu \pm c_3)$. We chose $c_.$ predefined values as 0.5, 0.025, and 0.15 for $\beta, \gamma$, and $\mu$ respectively. We then calculated the 95% HPD intervals for all the posterior distributions. Consequently, we calculated the percentage of HPD intervals that fell within the ROPE interval. Figure S14 shows the values that were calculated for the posterior distributions (under Assumption 2) of $\beta$, $\gamma$, and $\mu$ for sub-population 1. The percentages in ROPE for all the other sub-populations are included in a file on GitHub (the link is provided at the end of this document). Figure S16 illustrates the comparison of ROPE percentages and posterior medians for recovery rates.

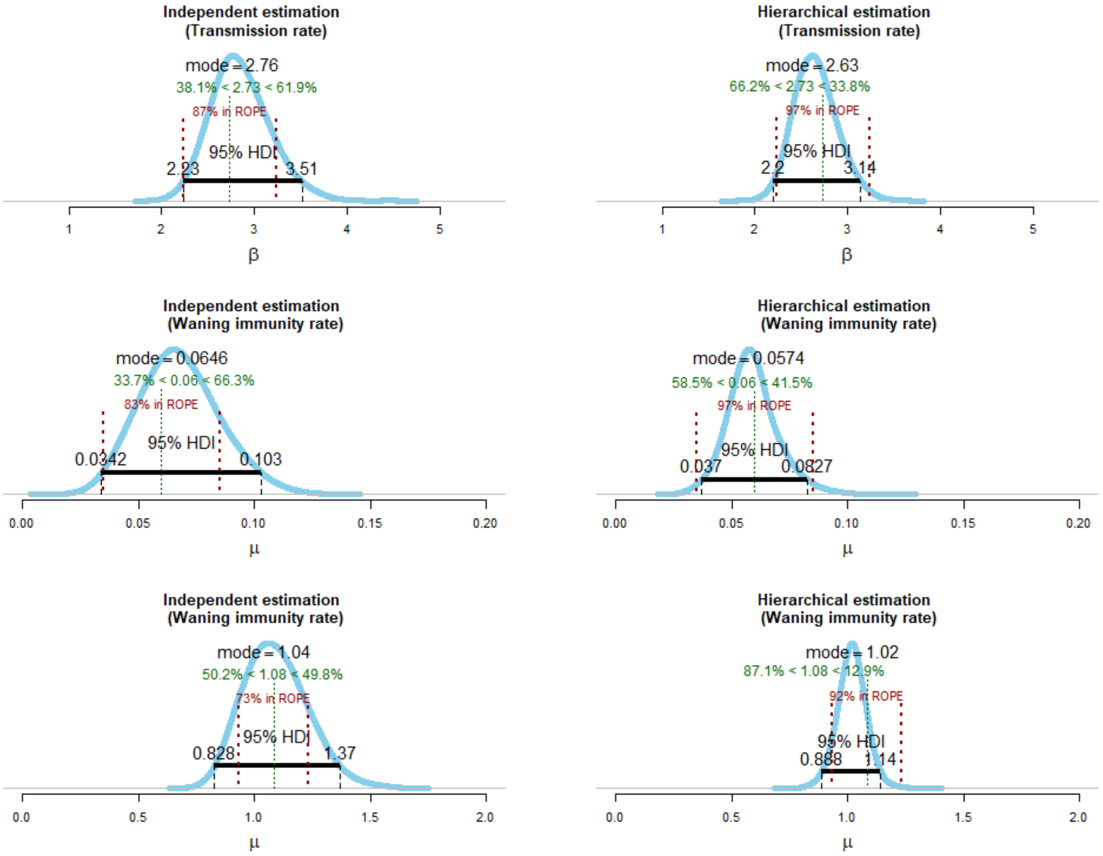


Figure S14: Percentages of ROPE for sub-population 1 for the dataset at $R_0 = 2.5$.

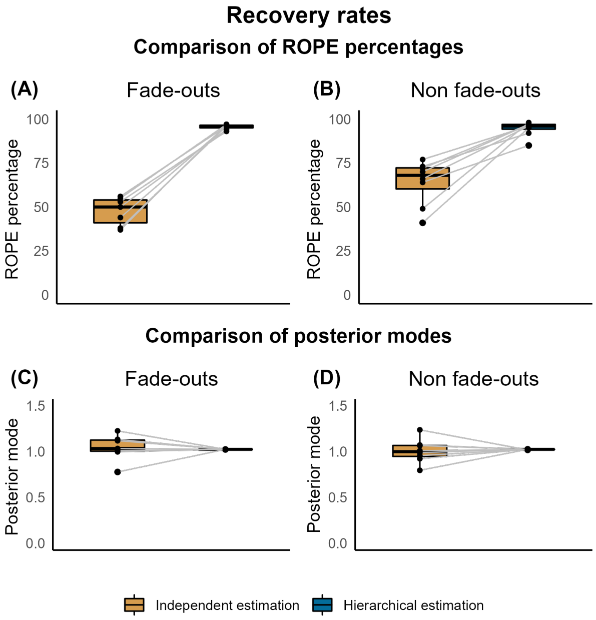


Figure S15: Comparison of ROPE percentages and posterior modes for recovery rates when epidemic fade-outs and non-fade-outs are observed. Dataset: at $R_0 = 2.5$

#### S2.7 Comparison of hyper-parameters under different assumptions

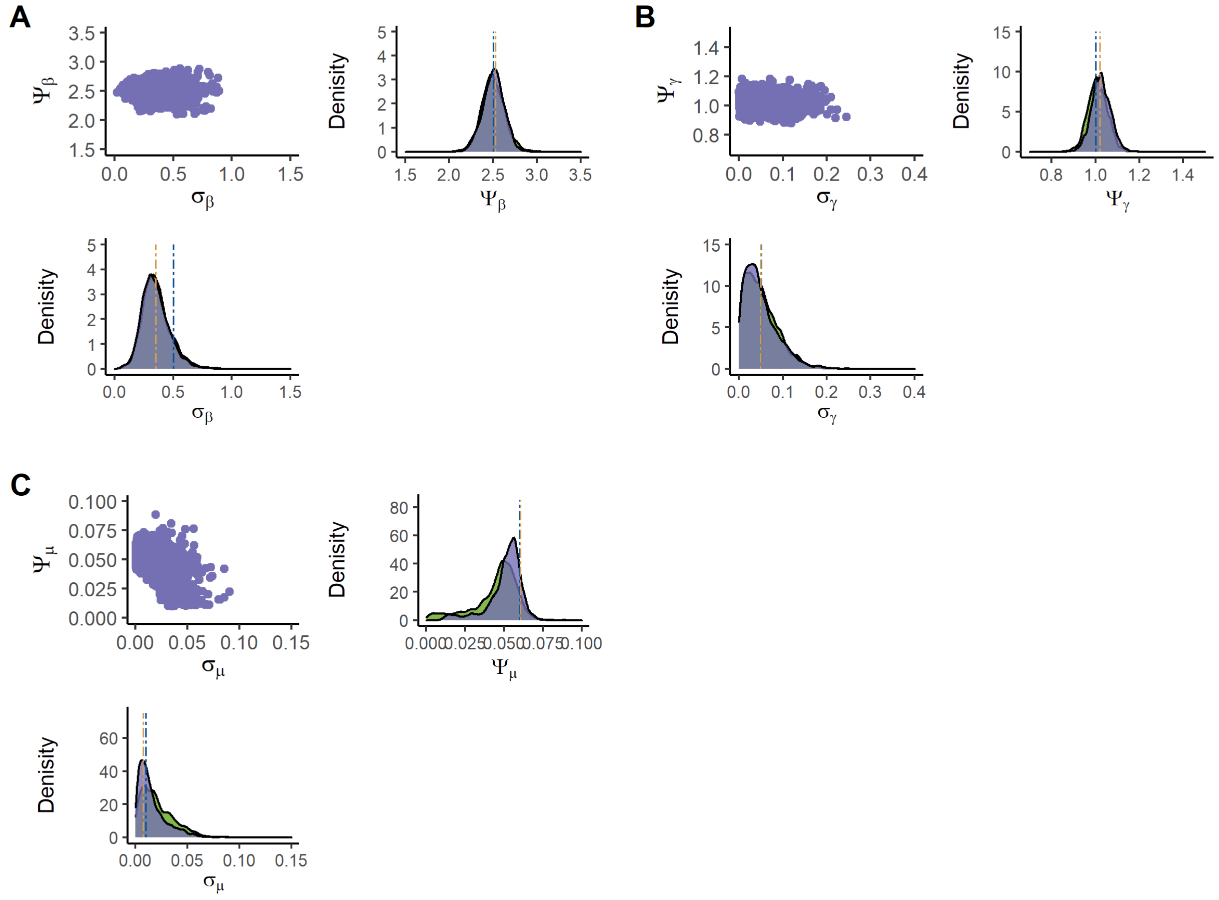


Figure S16: Marginal posterior distributions for the hyper-parameters for $\beta$ (A), $\gamma$ (B) and $\mu$ (C) for the datset at $R_0 = 2.5$. Purple : Posteriors under assumption 2 (prior excludes $\mu = 0$). Green: Posteriors under assumption 1 (prior admits $\mu = 0$). Blue dashed line: Parameter value. Orange dashed line: Mean of the sub-population specific parameters..

#### S2.8 Calculation of the epidemic fade-out probabilities

In Section 4.1 of the paper, we have calculated the epidemic fade-out probabilities of estimated parameters as follows.

Let $\boldsymbol{\theta}^*$ be a sample from the joint posterior distribution of the parameters of the $SIRS$ model. Given this sample, we first generate the endemic points $S(D), I(D)$ corresponding to the deterministic (ODE) SIRS system. Similar to Ballard, Bean, and Ross (2016), then the state, $S(t), I(t)$, of a stochastic SIRS system,

(Condition 1) Entered the first trough if $((S(t) < S(D))\&(I(t) <= I(D))$

(Condition 2) Left the first trough if $(I(t) >= 2I(D))$

(Condition 3) Left the first trough if $((S(t) >= S(D))\&(I(t) >= I(D)))$

(Condition 4) Disease extinction if $I(t) = 0$

Then we generated 1500 sample paths up to 35 days using the Gillespie algorithm with the same parameter set. If the state, $S(35), I(35)$ of a sample path satisfied Conditions 1 and 2 or Conditions 1 and 3, we defined that the sample path observed multiple outbreaks. If Conditions 1 and 4 were satisfied, we defined that the sample path observed an epidemic fade-out. The probability of epidemic fade-out for the estimated parameter set $\boldsymbol{\theta}^*$ was defined as the proportion of sample paths that observed epidemic fade-outs out of the sample paths that satisfied Condition 1 (i.e., paths that did not initially die-out).

#### S3 Identifying a major outbreak

At the hyper-parametric level, we considered the hyper means of the dataset, $(\psi_\beta, \psi_\gamma, \psi_\mu)$. Using this parameter set, we then generated a deterministic time plot of the $SIRS$ model and identified a time interval where the first trough would lie. We generally considered this time period to be where the deterministic path and the endemic line intersect. See panel (A) of Figure S17 for the dataset with hyper means $(\psi_\beta, \psi_\gamma, \psi_\mu) = (2.5, 1, 0.06)$. However, accounting for the stochastic effects of a stochastic $SIRS$ model, as well as the variability of the sub-population specific parameters, we also reduced the minimum number of days the stochastic system will enter the first trough to be less than that of the deterministic counterpart at the hyperparametric levels. This number for our original dataset with hyper means $(\psi_\beta, \psi_\gamma, \psi_\mu) = (2.5, 1, 0.06)$ is 8 days. See Table S8 for other datsets.

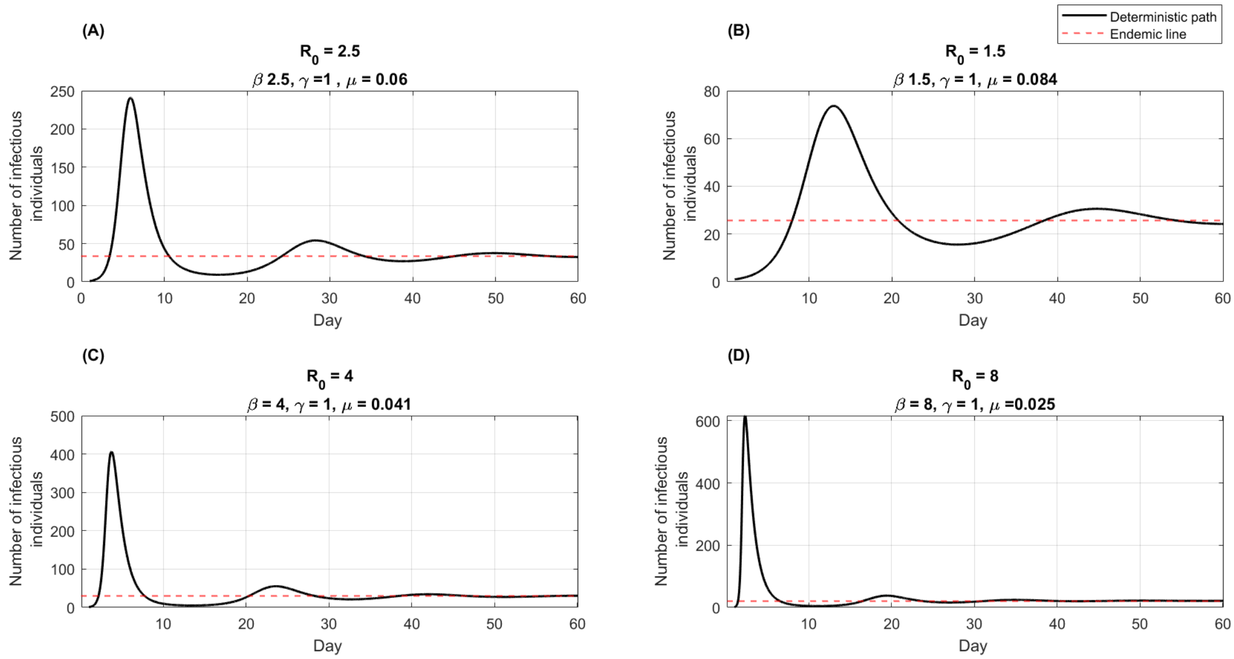


Figure S17: Deterministic paths (in black) of the $SIRS$ models considered for the four datasets with parameters as hyper means. Red dashed lines are the endemic lines.

Table S8: The time of entering the first trough under different $R_0$ values accounting for stochastic effects

| $R_0$ **at the hyperparametric level** | **Day the stochastic $SIRS$ model may enter the first trough** |
|---|---|
| 1.5 | 14 |
| 2.5 | 8 |
| 4 | 7 |
| 8 | 5 |

After reaching the first trough, if the stochastic system does not reach zero infectious individuals by the time it reaches day 35, and the number of infectious individuals is greater than 1, we consider it a non-fade-out. If the system has reached the first trough and the number of infectious individuals has reached zero during the time period of the trough, it is an epidemic fade-out.

#### S4 Performance of the estimation framework under different parameter regions

##### S4.1 Synthetic datasets

We generated parameter sets for each sub-population for the three new datasets from truncated normal distributions specified in Table S9. Similar to the data generation process we described in the main manuscript, we generated synthetic datasets as illustrated in Figures S18, S19 and S20. Each dataset consisted of 15 outbreaks. An outbreak in a sub-population started with one infectious individual and 999 susceptible individuals. The time period considered was 35 days.

Table S9: Sub-population specific parameters generation

| $R_0$ **at the hyperparametric level** | **Sub-population specific parameter generation** $\mathbf{TN}(a, b^2, c, d)$ | | |
|---|---|---|---|
| | $\beta$ | $\gamma$ | $\mu$ |
| 1.5 | $\text{TN}(1.5, 0.05^2, 1.25, 2.75)$ | $\text{TN}(1, 0.05^2, 0, 4)$ | $\text{TN}(0.084, 0.01^2, 0.0625, 1)$ |
| 4 | $\text{TN}(4, 0.25^2, 2.75, 6)$ | $\text{TN}(1, 0.05^2, 0, 4)$ | $\text{TN}(0.041, 0.01^2, 0.0330, 0.0625)$ |
| 8 | $\text{TN}(8, 0.25^2, 6, 12)$ | $\text{TN}(1, 0.05^2, 0, 4)$ | $\text{TN}(0.025, 0.01^2, 0.01, 0.0330)$ |

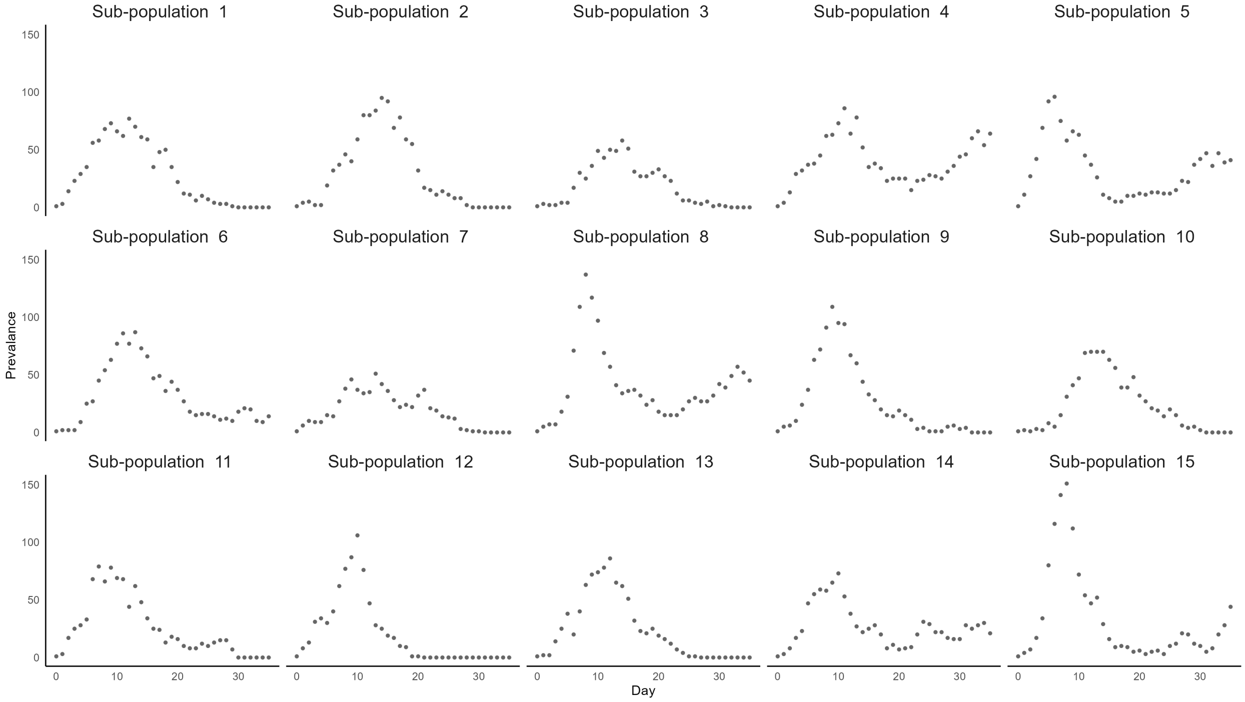


Figure S18: Synthetic dataset with $R_0 = 1.5$ at the hyper-parametric level.

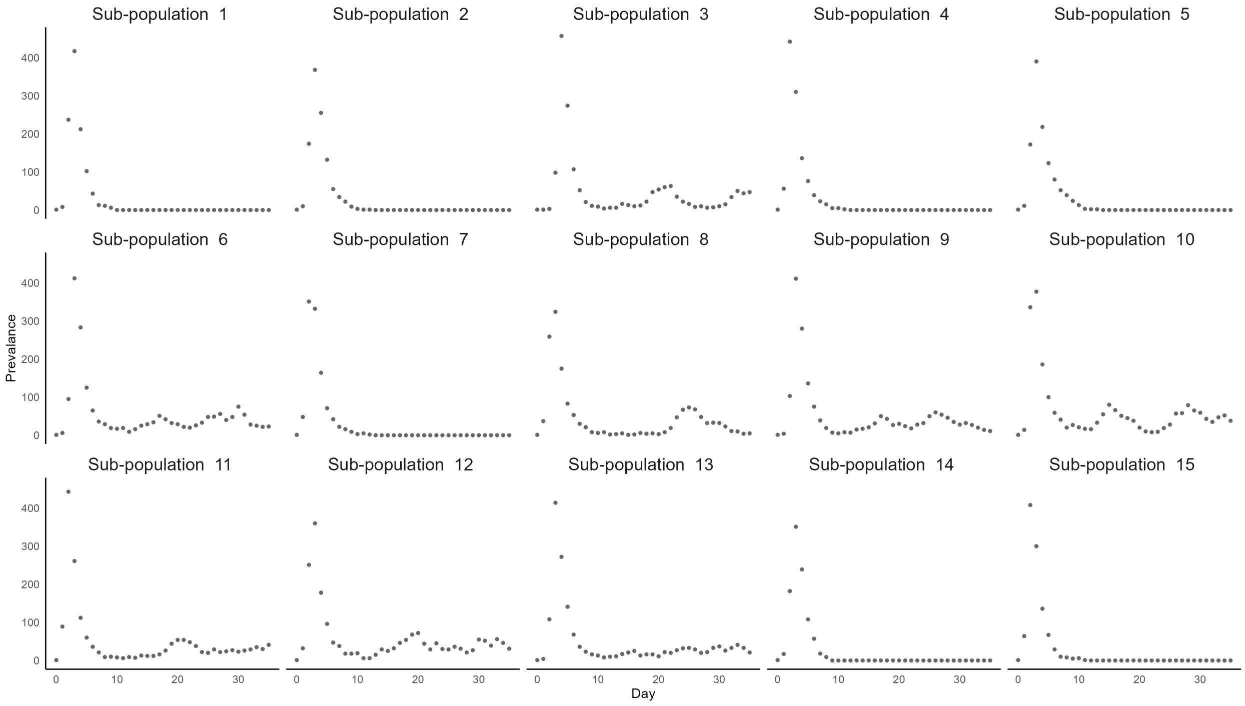


Figure S19: Synthetic dataset with $R_0 = 4$ at the hyper-parametric level.

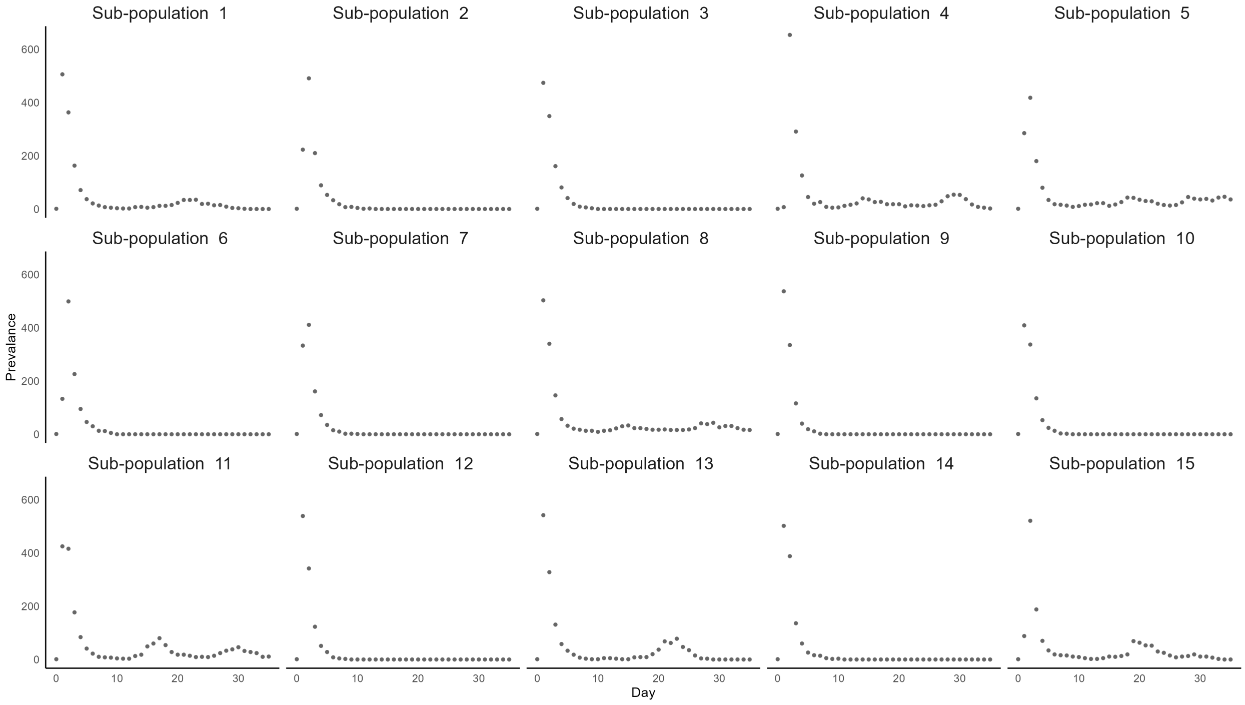


Figure S20: Synthetic dataset with $R_0 = 8$ at the hyper-parametric level.

##### S4.2 Posterior distributions under independent vs. hierarchical estimation

**Dataset 1:** $R_0 = 1.5$

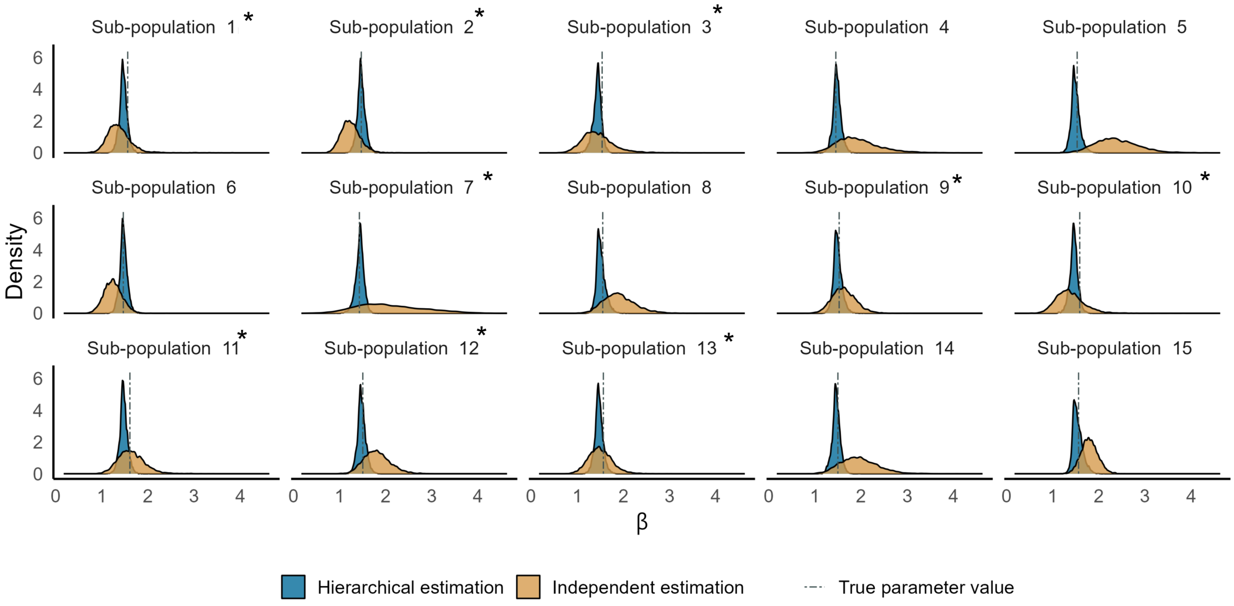


Figure S21: Posterior distribution for $\beta$ for the dataset with $R_0 = 1.5$ at the hyperparametric level with assumption 1 (prior admits $\mu = 0$). Asterisks represent the sub-populations that experienced a fade-out. For each sub-population, the posterior distributions under an independent (orange) and hierarchical (blue) estimation framework are overlaid. The blue dashed lines are the true parameter values.

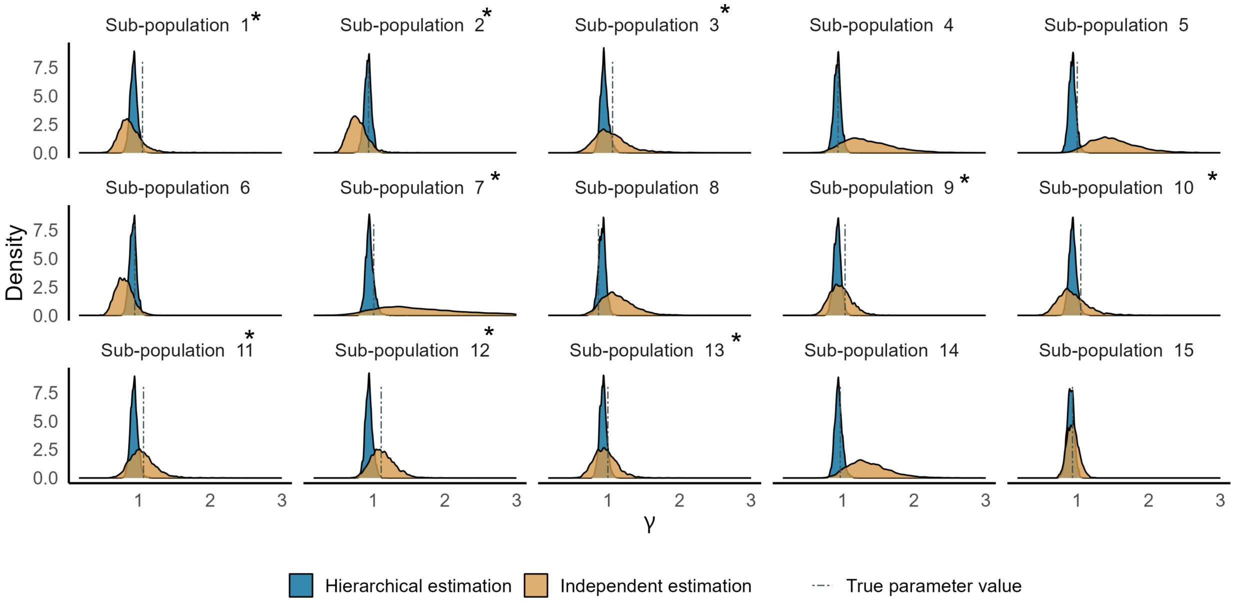


Figure S22: Posterior distribution for $\gamma$ for the dataset with $R_0 = 1.5$ at the hyperparametric level with assumption 1 (prior admits $\mu = 0$). Asterisks represent the sub-populations that experienced a fade-out. For each sub-population, the posterior distributions under an independent (orange) and hierarchical (blue) estimation framework are overlaid. The blue dashed lines are the true parameter values.

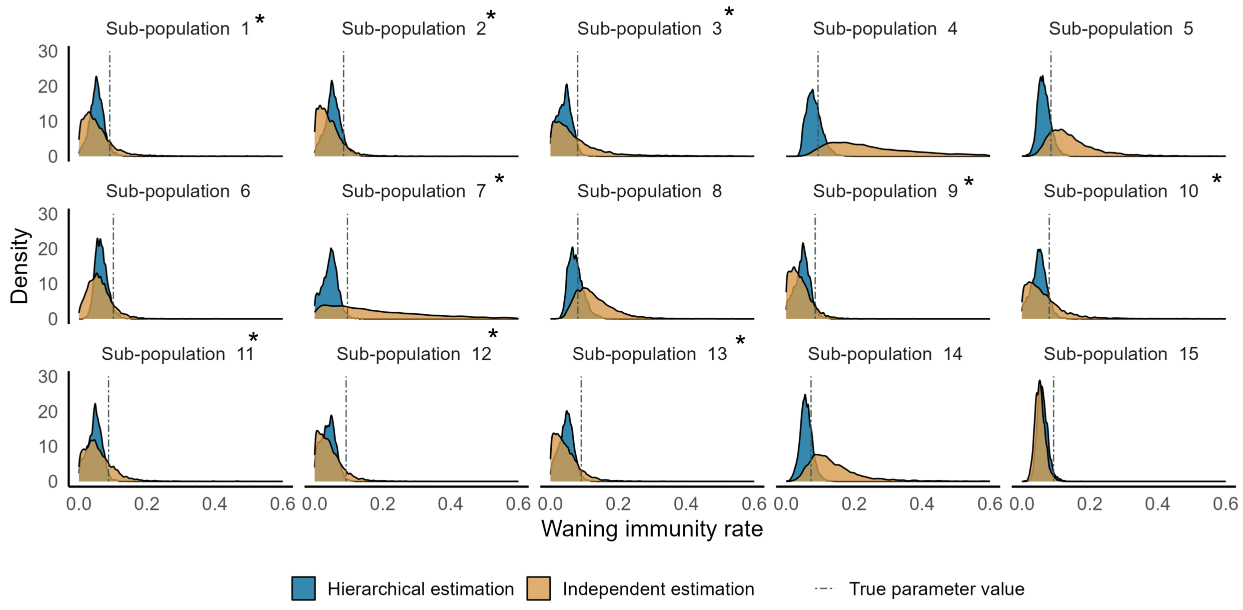


Figure S23: Posterior distribution for $\mu$ for the dataset with $R_0 = 1.5$ at the hyperparametric level with assumption 1 (prior admits $\mu = 0$). Asterisks represent the sub-populations that experienced a fade-out. For each sub-population, the posterior distributions under an independent (orange) and hierarchical (blue) estimation framework are overlaid. The blue dashed lines are the true parameter values.

**Dataset 2:** $R_0 = 4$

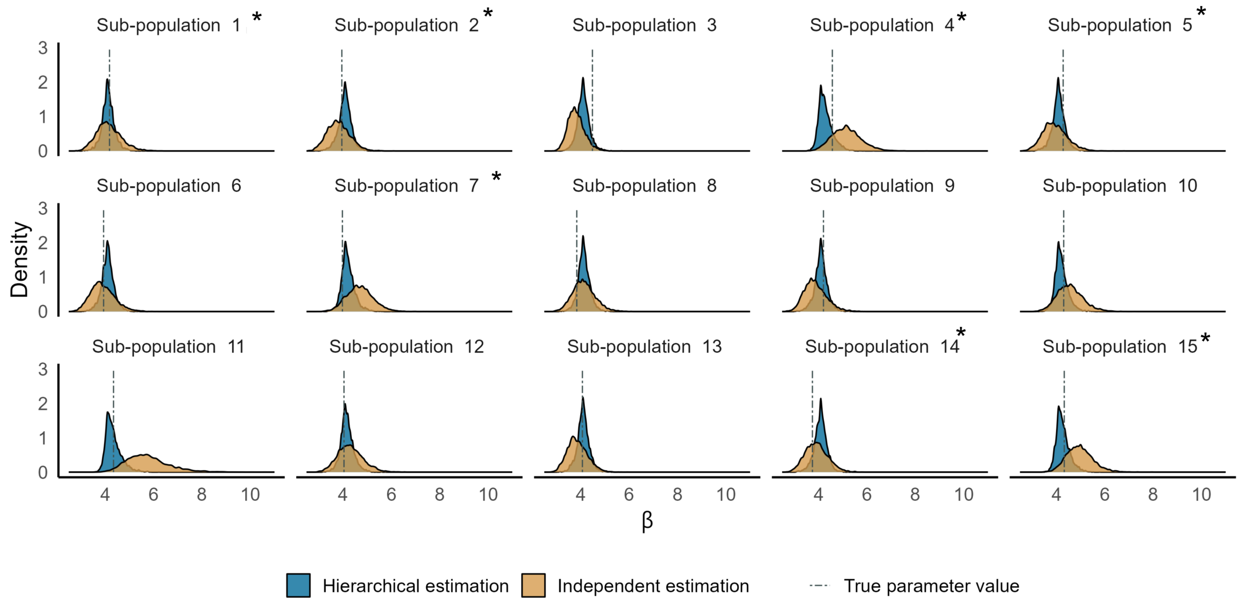


Figure S24: Posterior distribution for $\beta$ for the dataset with $R_0 = 4$ at the hyperparametric level with assumption 1 (prior admits $\mu = 0$). Asterisks represent the sub-populations that experienced a fade-out. For each sub-population, the posterior distributions under an independent (orange) and hierarchical (blue) estimation framework are overlaid. The blue dashed lines are the true parameter values.

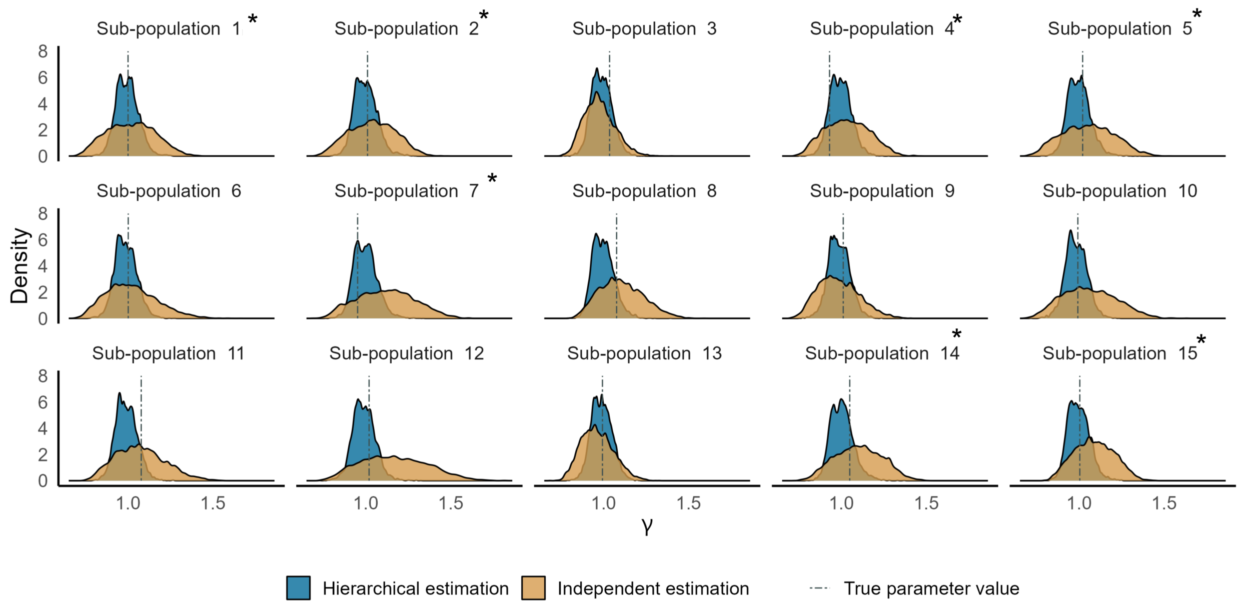


Figure S25: Posterior distribution for $\gamma$ for the dataset with $R_0 = 4$ at the hyperparametric level with assumption 1 (prior admits $\mu = 0$). Asterisks represent the sub-populations that experienced a fade-out. For each sub-population, the posterior distributions under an independent (orange) and hierarchical (blue) estimation framework are overlaid. The blue dashed lines are the true parameter values.

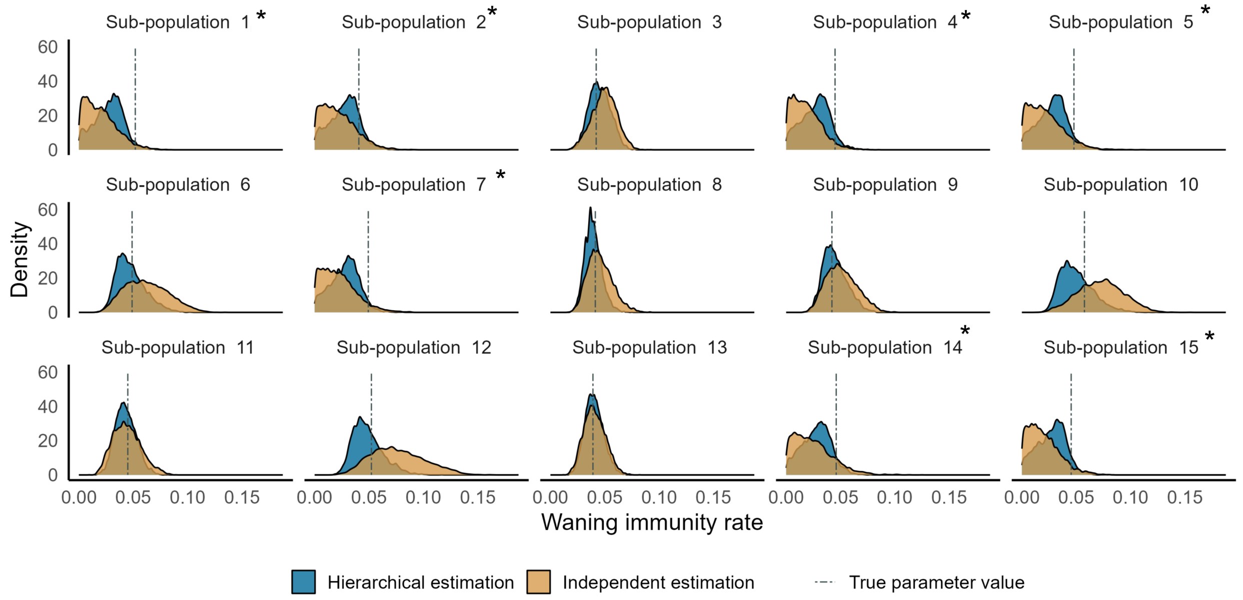


Figure S26: Posterior distribution for $\mu$ for the dataset with $R_0 = 4$ at the hyperparametric level with assumption 1 (prior admits $\mu = 0$). Asterisks represent the sub-populations that experienced a fade-out. For each sub-population, the posterior distributions under an independent (orange) and hierarchical (blue) estimation framework are overlaid. The blue dashed lines are the true parameter values.

**Dataset 3:** $R_0 = 8$

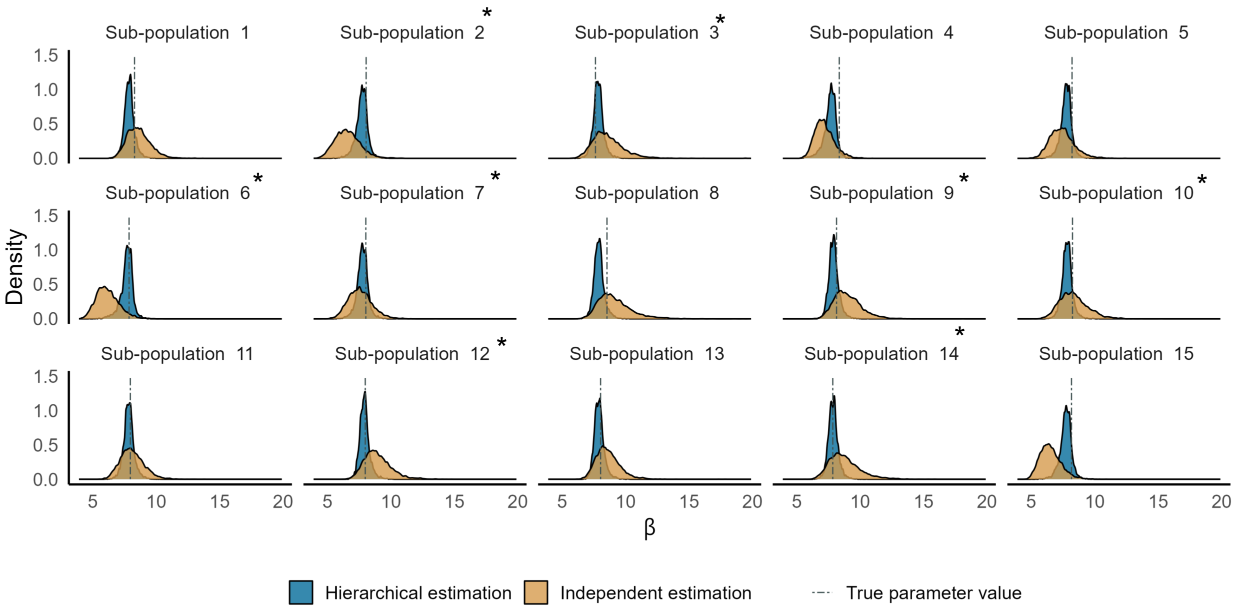


Figure S27: Posterior distribution for $\beta$ for the dataset with $R_0 = 8$ at the hyperparametric level with assumption 1 (prior admits $\mu = 0$). Asterisks represent the sub-populations that experienced a fade-out. For each sub-population, the posterior distributions under an independent (orange) and hierarchical (blue) estimation framework are overlaid. The blue dashed lines are the true parameter values.

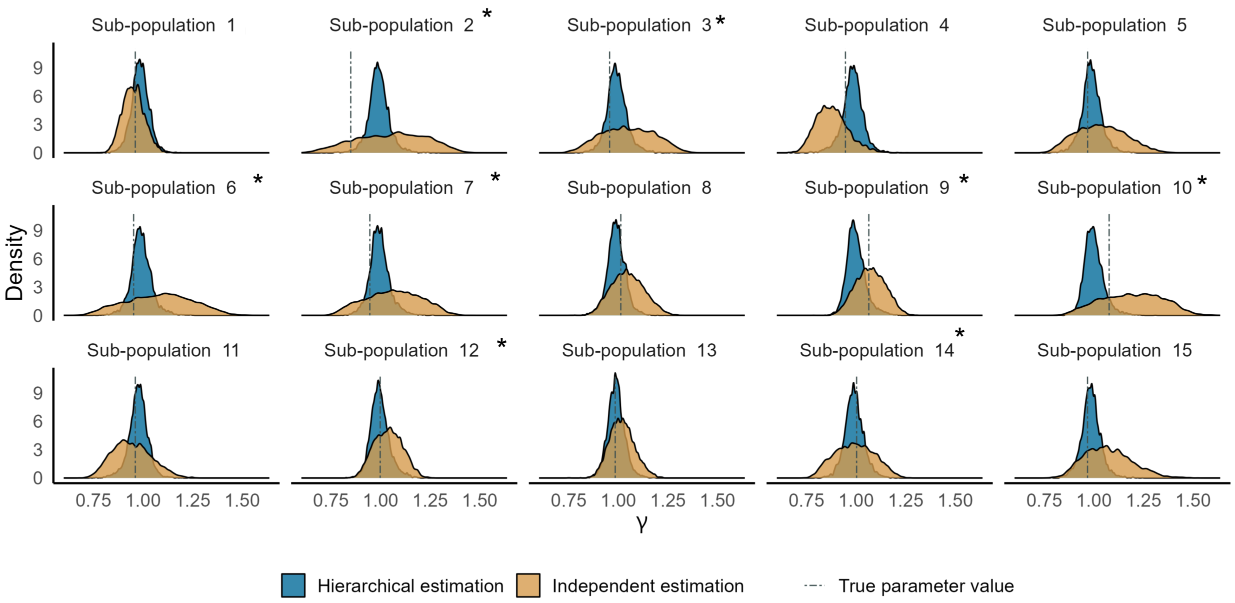


Figure S28: Posterior distribution for $\gamma$ for the dataset with $R_0 = 8$ at the hyperparametric level with assumption 1 (prior admits $\mu = 0$). Asterisks represent the sub-populations that experienced a fade-out. For each sub-population, the posterior distributions under an independent (orange) and hierarchical (blue) estimation framework are overlaid. The blue dashed lines are the true parameter values.

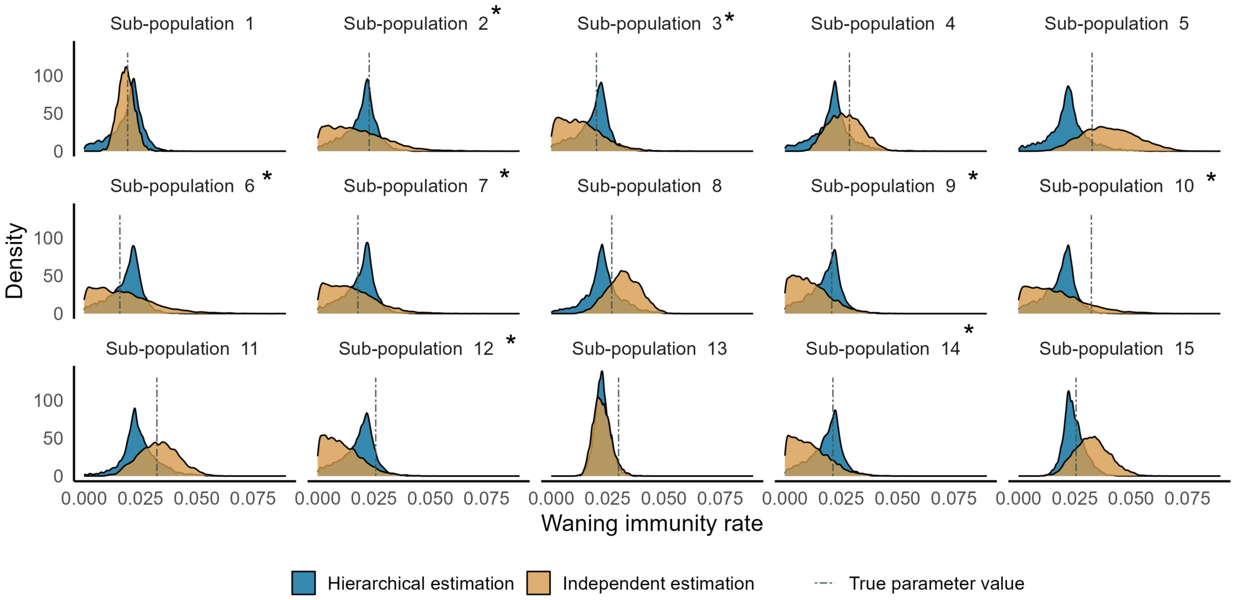


Figure S29: Posterior distribution for $\mu$ for the dataset with $R_0 = 8$ at the hyperparametric level with assumption 1 (prior admits $\mu = 0$). Asterisks represent the sub-populations that experienced a fade-out. For each sub-population, the posterior distributions under an independent (orange) and hierarchical (blue) estimation framework are overlaid. The blue dashed lines are the true parameter values.

#### S4.3 Hyper parameters

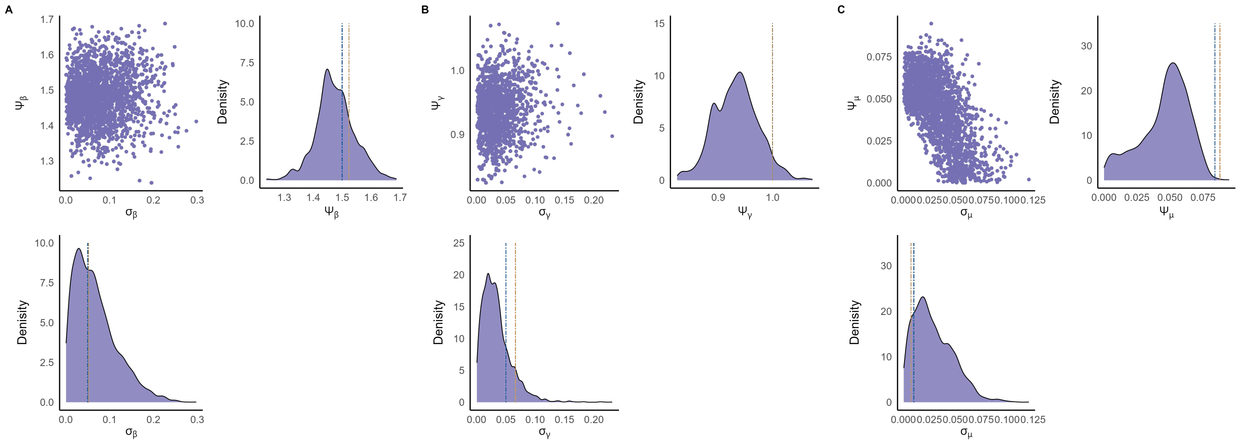


Figure S30: Posterior distributions of the hyperparameters for the dataset with $R_0 = 1.5$ at the hyperparametric level with assumption 1 (prior admits $\mu = 0$). Asterisks represent the sub-populations that experienced a fade-out. For each sub-population, the posterior distributions under an independent (orange) and hierarchical (blue) estimation framework are overlaid. The blue dashed lines are the true parameter values.

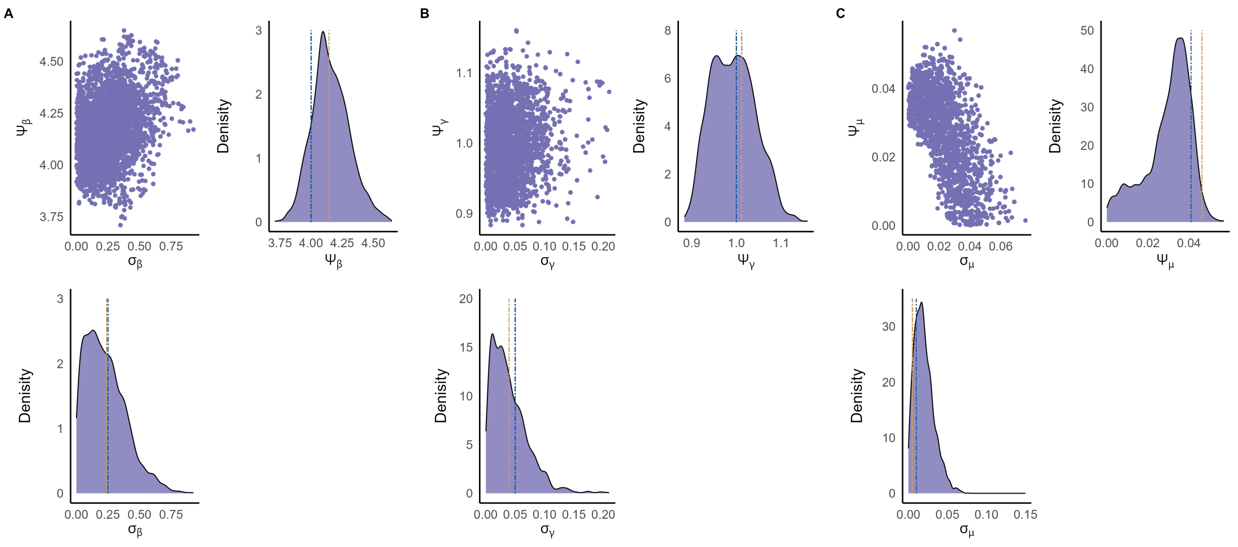


Figure S31: Posterior distributions of the hyperparameters for the dataset with $R_0 = 4$ at the hyperparametric level with assumption 1 (prior admits $\mu = 0$). Asterisks represent the sub-populations that experienced a fade-out. For each sub-population, the posterior distributions under an independent (orange) and hierarchical (blue) estimation framework are overlaid. The blue dashed lines are the true parameter values.

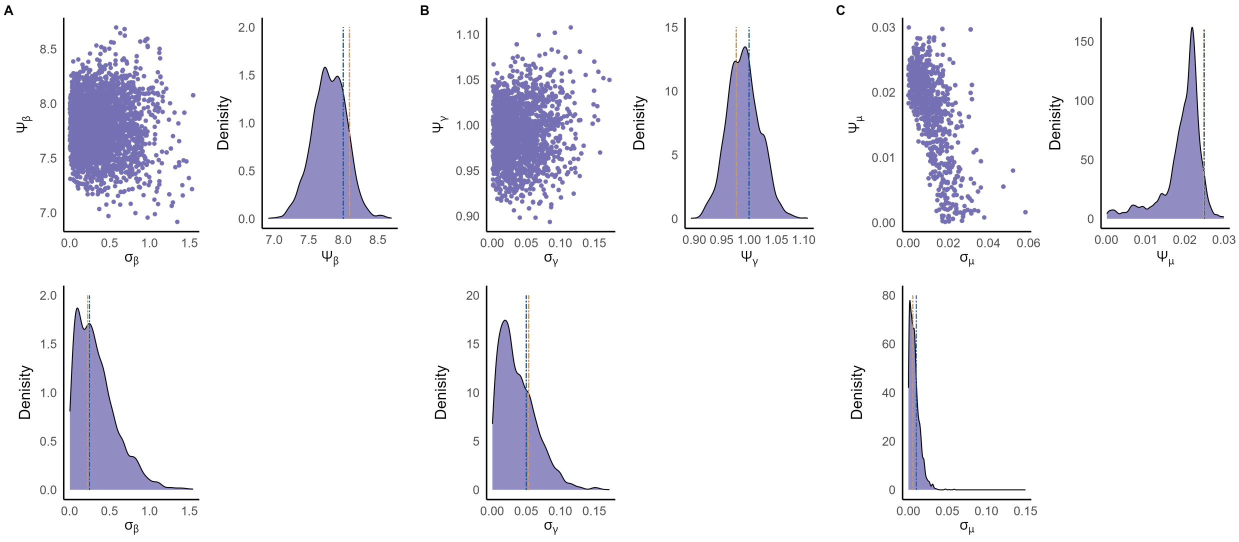


Figure S32: Posterior distributions of the hyperparameters for the dataset with $R_0 = 8$ at the hyperparametric level with assumption 1 (prior admits $\mu = 0$). Asterisks represent the sub-populations that experienced a fade-out. For each sub-population, the posterior distributions under an independent (orange) and hierarchical (blue) estimation framework are overlaid. The blue dashed lines are the true parameter values.

##### S4.4 ROPE calculation

We calculated each dataset's ROPE percentages for $\mu$ with $(\psi_\mu \pm c)$. $c$ value for datasets 1, 2, and 3 were 0.05, 0.025, and 0.008. The calculated ROPE percentages are included in the GitHub folder.

#### S5 MATLAB Codes

The codes can be found on GitHub at: https://github.com/PunyaAlahakoon/waning_immunity_stochastic_SIRS.git
